# Investigating the relationship between the immune response and the severity of COVID-19: a large-cohort retrospective study

**DOI:** 10.1101/2024.06.20.24309246

**Authors:** Riccardo Giuseppe Margiotta, Emanuela Sozio, Fabio Del Ben, Antonio Paolo Beltrami, Daniela Cesselli, Martina Fabris, Francesco Curcio, Carlo Tascini, Guido Sanguinetti

## Abstract

The COVID-19 pandemic has left an indelible mark globally, presenting numerous challenges to public health. This crisis, while disruptive and impactful, has provided a unique opportunity to gather precious clinical data extensively. In this observational, case-control study, we utilized data collected at the Azienda Sanitaria Universitaria Friuli Centrale, Italy, to comprehensively characterize the immuno-inflammatory features in COVID-19 patients. Specifically, we employed multicolor flow cytometry, cytokine assays, and inflammatory biomarkers to elucidate the interplay between the infectious agent and the host’s immune status. We characterized immuno-inflammatory profiles within the first 72 hours of hospital admission, stratified by age, disease severity, and time elapsed since symptom onset. Our findings indicate that patients admitted to the hospital shortly after symptom onset exhibit a distinct pattern compared to those who arrive later, characterized by a more active immune response and heightened cytokine activity, but lower markers of tissue damage. We used univariate and multivariate logistic regression models to identify informative markers for outcome severity. Predictors incorporating the immuno-inflammatory features significantly outperformed standard baselines, identifying up to 59% of patients with positive outcomes while maintaining a false omission rate as low as 4%. Overall, our study sheds light on the immuno-inflammatory aspects observed in COVID-19 patients prior to vaccination, providing insights for guiding the clinical management of first-time infections by a novel virus.

## 1 Introduction

The COVID-19 pandemic had a devastating impact worldwide, causing significant morbidity and mortality. The strain on healthcare systems has been particularly pronounced, with hospitals reaching their capacity, especially for intensive care. Notably, the virus has disproportionately affected certain demographic groups, revealing stark disparities in health outcomes [1]. The reasons for these disparities are multifaceted, encompassing socioeconomic and genetic factors, pre-existing health conditions, and, potentially, differences in immune responses. While our comprehension of the complex interplay between SARS-CoV-2 and the human immune system has improved over the years, the picture is still incomplete. During the early phase of the pandemic, it became evident that the severity of COVID-19 was influenced not exclusively by the virus but also by the host’s immune response. An overactive or dysregulated response in some individuals led to severe disease and even death, while others exhibited a more controlled response leading to a better outcome [2, 3]. The mechanisms governing these varied responses remain incompletely understood, emphasizing the need for in-depth studies on the interaction between the virus and the immune system.

Understanding the immune-inflammatory response to COVID-19 is also crucial for gaining insights into related diseases. One important example is *sepsis*, a severe condition that can be caused by bacteria, fungi, or viruses, which currently lacks a specific treatment [4]. COVID-19 hospitalized patients should intrinsically be regarded as septic: the majority of critically ill patients (78%) met Sepsis-3 criteria for septic shock with acute respiratory distress syndrome (ARDS) as the most frequent organ dysfunction (88%) [5, 6]. Recent advances in understanding sepsis have led to the belief that the majority of sepsis-related deaths are not caused by the initial hyperinflammatory state, but rather by the suppression of the immune system, known as sepsis-induced immunoparalysis [7]. One of the mainstays of treatment for severely ill COVID-19 patients on supplemental oxygen has been glucocorticoids, which are anti-inflammatory and immunosuppressive drugs [8]. However, for some COVID-19 and septic patients, immunostimulation rather than immunosuppression can be a more appropriate approach: understanding the underlying mechanisms of disease progression is essential to prevent inappropriate treatments. Furthermore, a deeper understanding of the immune response to SARS-CoV-2 could aid in developing predictive tools to identify individuals at risk of severe outcomes. This would be extremely important for prioritizing hospitalizations and allocating healthcare resources. Clinical scores have been utilized as a method for predicting outcomes and stratifying risks, such as the 4C Mortality Score, and they have shown promising results [9, 10].

Throughout the pandemic, our hospital, the *Ospedale Santa Maria della Misericordia* of Udine (Italy), has witnessed a large number of hospitalizations among COVID-19 patients, presenting an unprecedented opportunity to accumulate extensive information about a single disease. We therefore used the collected clinical data to create a comprehensive retrospective database, the MANDI registry (“MAnagement coroNavirus Disease In hospital registry” – authorization of DG, decree n. 957, 10/09/2021). This database has allowed us to delve into the study of immune status and identify mid-regional pro-adrenomedullin (proADM) as an effective biomarker for predicting outcomes, in association with lactate dehydrogenase (LDH) and C-reactive protein (CRP) [11, 12]. Additionally, we have investigated the role of cytokines in the setting of COVID-19-related pericarditis [13] and employed machine learning techniques to develop predictive tools while deepening our understanding of cytokines and serum proteomics [14, 15]. Building upon these achievements, in the present paper we aim to provide a more comprehensive study of the immuno-inflammatory profiles of COVID-19 patients. Our focus is on analyzing the distribution of monocyte and lymphocyte populations observed during the infection, utilizing immunological data obtained by flow cytometry. Additionally, our analysis includes serological biomarkers (CRP, proADM, and LDH), as well as cytokines, thus covering different aspects of the host’s immuno-inflammatory response. We believe that a better comprehension of the pathophysiology of COVID-19 could provide insights into the broader management of sepsis. The contribution of this study is twofold. Firstly, we present a detailed phenotypic characterization of COVID-19 patients, with an emphasis on viral etiology, shedding light on the specific immune cell profiles associated with the infection. Secondly, we conduct a predictive analysis to identify the most informative biomarkers for predicting patient outcomes, using several clinical scores as baselines for comparison.

The rest of the manuscript is organized as follows: in Sec. 2, we characterize the patients’ immune-inflammatory response through descriptive statistics. Sec. 3 presents our analysis aimed at predicting patient outcomes. Given the multifaceted nature of our analyses across different data types, we discuss the results within their respective sections as they are presented. Additionally, we provide a comprehensive summary of the main findings and a broader discussion in Sec. 4. Finally, a detailed description of the materials and methods employed in our study is provided in Sec. 5.

## 2 Immuno-inflammatory profiles of COVID-19 patients

The data collection process was conducted between March 2020 and April 2021, covering the first three waves of the pandemic. The resulting data comprises approximately 900 records and includes a variety of variables collected upon hospital admission. These variables encompass demographic information, individual comorbidities, monocyte and lymphocyte counts, and, for smaller subsets of patients, measurements of cytokines and serological biomarkers. Crucially, none of the patients had been vaccinated against SARS-CoV-2 yet. Below is a complete list of all the immuno-inflammatory features considered in our analysis, with the abbreviations used throughout this manuscript:

- **flow cytometry** (FC set): counts of white blood cells (WBC), monocytes (Mono), lymphocytes (Lymph), T and B cells (CD3, CD4, CD8, CD19), natural killer cells (NK), recent thymic emigrants (RTE), the percentage of monocytes, CD4, and CD8 positive cells expressing HLA-DR (HLA^+^_%Mono_, HLA^+^_%CD4_, HLA^+^_%CD8_), the percentage of RTE-CD4 cells (RTE _%CD4_), and the monocytes HLA-DR mean intensity fluorescence (HLA^+^IF _Mono_).
- **cytokines** (CK set): interleukin 10 (IL10), IL1-*β* (IL1B), sIL2R-*α*/sCD25 (IL2R), interleukin 6 (IL6), interleukin 8 (IL8), chemokine IP10/CXCL10 (IP10), and interferon-*γ* (INF-*γ*).
- **biomarkers** (BM set): mid-regional pro-adrenomedullin (proADM), a marker of endothelial response to inflammation and tissue damage, lactate dehydrogenase (LDH), a marker of cell proliferation and/or damage, and C-reactive protein (CRP).

We also collected flow cytometry and demographic data from approximately 370 asymptomatic outpatients, designated as the ***control set*** in our analysis. Importantly, the control set exhibits matching statistics with a smaller cohort of healthy individuals (approximately 90 cases), as depicted in the Supp. Fig. 8. For hospitalized patients, we collected information on patients’ outcomes, including survival status, as well as the treatments they received. Additionally, we assessed disease severity with a 4-point ordinal scale following the World Health Organization’s guidance [16], which we refer to as the WHO scale.

Contingent upon data availability, we utilized the Charlson Comorbidity Index and four clinical indices as baseline scores to evaluate patients’ statuses at admission to the hospital, as listed below:

- **CCI** (Charlson Comorbidity Index [17]): not conventionally a score, it is used to predict the ten-year mortality for a patient who may have a range of comorbid conditions. Each condition is assigned a score of 1, 2, 3, or 6, depending on the risk of dying associated with each one.
- **SOFA** (Sequential Organ Failure Assessment [18]): involves six organ systems (respiratory, cardiovascular, hepatic, coagulation, renal, and neurological), and it is used for mortality prediction of intensive care unit patients [19] and for the diagnosis of sepsis [18].
- **NEWS** (National Early Warning Score [20]): determines the degree of illness of a patient and prompts critical care intervention [21] evaluating respiration rate, oxygen saturation, systolic blood pressure, pulse rate, level of consciousness, and temperature.
- **qCSI** (Quick COVID-19 Severity Index [22]): based on respiratory rate, pulse oximetry, and speech evaluation, it is used to predict the 24-hr risk of critical respiratory illness in COVID-19 patients.
- **4C** (Coronavirus Clinical Characterisation Consortium mortality score [23]): based on age, sex, number of comorbidities, respiratory rate, peripheral oxygen saturation, level of consciousness, urea level, and CRP, it is used for predicting the in-hospital mortality of patients admitted with COVID-19 [24, 25].

In the following sections, we provide a comprehensive description of the flow cytometry, cytokines, and biomarkers variables, by analyzing their relationships with the developed illness severity (WHO scale), the patient age, and the number of days elapsed between symptoms onset and hospitalization (Δ*t*_ons_). We remark that Δ*t*_ons_ is an anamnestic variable with some limitations since it relies on the ability of the patient to detect the symptoms and remember when they first occurred. For our analysis, we considered only patients aged between 30 and 100 and with Δ*t*_ons_ in the range 0 to 30 days, excluding those with a CCI greater than 6 to minimize the impact of confounding factors on the outcomes. The resulting records, grouped according to data availability of the three sets of features (FC, CK, BM), exhibit homogeneous characteristics, as illustrated in Table 1, with the exception of sample size, which is maximal for the FC set. Note, however, that outpatients are typically younger and have a larger proportion of females. For more information on the data collection process and preprocessing, we refer to Sec. 5.

**Table 1:** Demographics and clinical characteristics, for all records (FC set), and subsets of the FC see containing additional information on cytokines (CK set) and serological biomarkers (BM set). Only records with less than 50% of missing data (NANs) were included in each set. Dichotomous variables are presented as percentages of available data. Numerical variables are described by the median (Q_2_), first (Q_1_), and third quartile (Q_3_). The cutoff of the scores are those identified in the literature as pathological. A dash indicates missing information.

| outpatients |  |  |  |  |  |  |
| --- | --- | --- | --- | --- | --- | --- |
| set | N | sex (female) | age [Q<sub>2</sub> (Q<sub>1</sub>-Q<sub>3</sub>)] | $\Delta t_{\text{ons}}$ [Q<sub>2</sub> (Q<sub>1</sub>-Q<sub>3</sub>)] | CCI<2 | NANs |
| FC | 367 | 58.3% | 51 (36-61) | - | - | 22.1% |
| inpatients |  |  |  |  |  |  |
| set | N | sex (female) | age [Q<sub>2</sub> (Q<sub>1</sub>-Q<sub>3</sub>)] | $\Delta t_{\text{ons}}$ [Q<sub>2</sub> (Q<sub>1</sub>-Q<sub>3</sub>)] | CCI<2 | NANs |
| FC | 788 | 33.9% | 68 (58-76) | 9 [7-12] | 20.3% | 3.8% |
| CK | 297 | 34.3% | 67 (56-77) | 9 [7-12] | 22.7% | 0.9% |
| BM | 539 | 35.1% | 68 (58-76) | 9 [7-12] | 20.5% | 6.9% |
| set | SOFA>1 | NEWS>4 | qCSI>6 | 4C>8 | WHO>2 | OTI+death |
| FC | 63.0% | 28.3% | 2.0% | 48.7% | 63.2% | 24.1% |
| CK | 61.4% | 22.8% | 0.7% | 46.5% | 58.0% | 23.6% |
| BM | 60.5% | 24.0% | 1.7% | 51.0% | 57.9% | 20.8% |

### 2.1 Relationship between immune response and disease severity

We explored how the distributions of flow cytometry measurements, cytokine levels, and biomarker concentrations changed in relation to the disease severity observed during patients’ hospitalization. We used the WHO scale for this purpose, which categorizes disease severity as mild (1), moderate (2), severe (3), and critical (4); for more details, we refer to Sec. 5. The results are summarized in Fig. 1, where panel **a** displays the flow cytometry features. We observe that the average white blood cell count (WBC) displays a non-monotonic behavior: it is lower for inpatients with mild severity compared to outpatients (control), and it increases with the disease severity for inpatients. Interestingly, the average number of monocytes (Mono) remains relatively constant across the WHO levels, while their activation (HLA^+^IF_Mono_) shows a drastic reduction as severity increases. Similarly, a significant decrease in lymphocyte counts is observed both at the aggregate level (Lymph) and in specific subpopulations such as CD4, CD8, and NK cells. This implies that the observed increase in WBC is due to the populations of granulocytes. Notably, CD19 counts are lower among inpatients compared to the control set of outpatients, though variations across different WHO levels are not significant.

**Figure 1:**
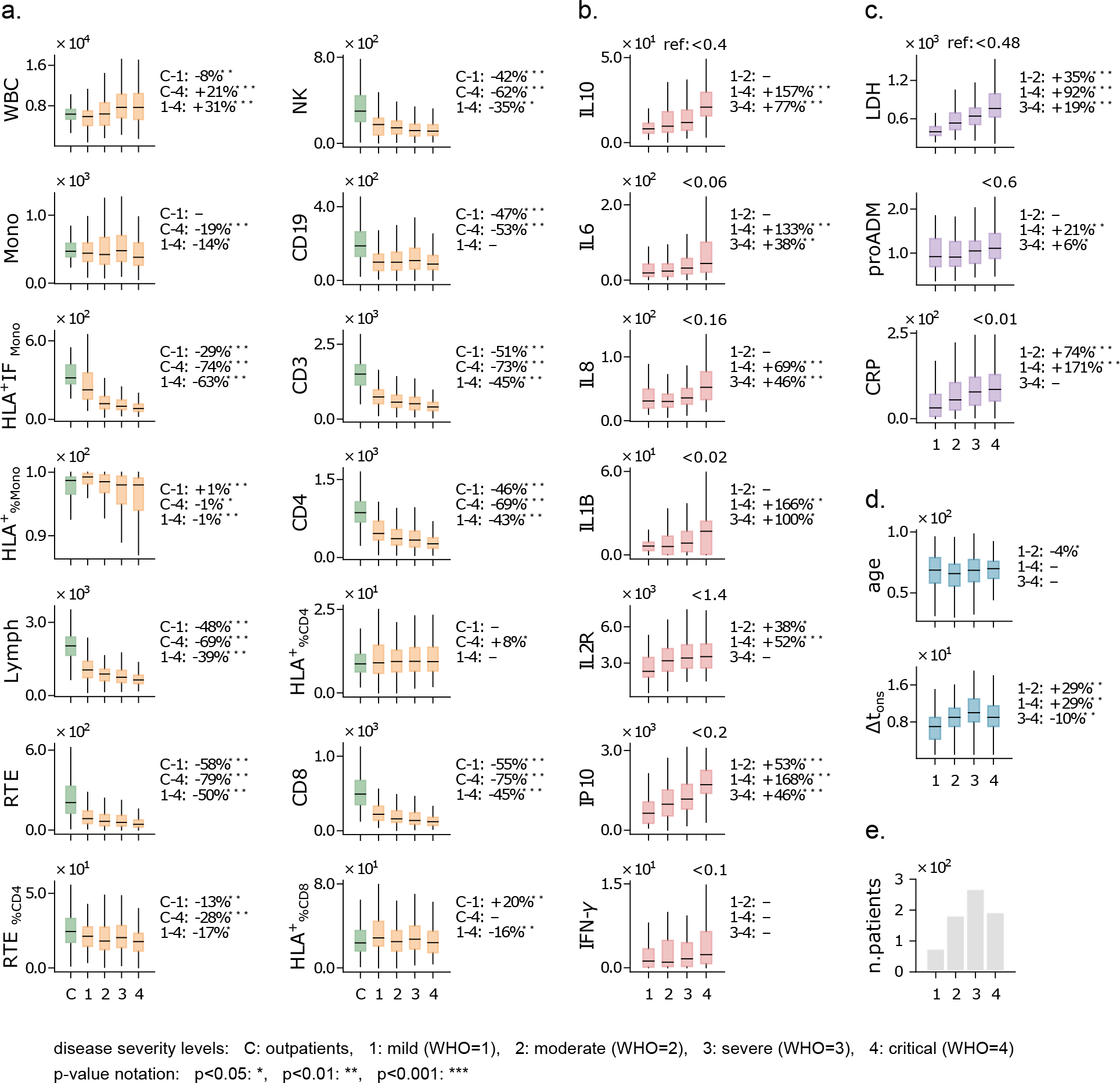
Feature distributions against disease severity. **a**) Boxplots of flow cytometry variables for patients stratified according to the WHO scale. Boxes include first-to-third quartiles, and the horizontal bar represents the median. The %-change of the median between severity levels is shown on the right of each plot (e.g. the WBC median is 8% lower in patients with mild disease (1) compared to outpatients (C)). **b**–**d**) Similar to panel **a**, for cytokines, biomarkers, age, and Δ*t*_ons_. Reference values for healthy individuals are shown at the top-right of each plot. **e**) Number of hospitalized patients across WHO levels. Measurement units: WBC and related subpopulations: U*/µ*L; HLA^+^IF_Mono_: U; cytokines: pg*/*mL; LDH: U*/*L; proADM: nmol*/*L; CRP: mg*/*L.

All cytokines are significantly overexpressed compared to the reference values, showing an increasing relationship with the WHO severity levels (Fig. 1-**b**). This pattern is especially prominent for IP10, a pro-inflammatory chemokine involved in recruiting immune cells to the infection site, and for IL10, an antiinflammatory cytokine that modulates immune responses and prevents excessive damage. A strong increase with disease severity is also observed for IL1B, a cytokine that plays a pivotal role in the overall response to infections, and IL6, which has a key role in the pro-inflammatory process as the major inducer of most acute phase reactants and is closely related to CRP [26, 27]. Recent studies have shown that patients with severe forms of COVID-19 have lower levels of interferon-gamma (IFN-*γ*) compared to patients with mild forms [28, 29]. In our case, instead, IFN-*γ* shows a moderate (non-significant) increase with the WHO severity levels. Overall, the increase in cytokine levels aligns with the well-documented phenomenon of the “cytokine storm” observed in COVID-19 patients, a dysregulation characterized by heightened cytokine levels associated with pro-inflammatory signals (IL6, IL8, and IP10) as well as immunomodulatory ones (IL10 and IL2R) [30, 31].

Similarly to cytokine levels, all analyzed biomarkers exhibit high concentrations and an increasing trend with the WHO severity level (Fig. 1-**c**). This trend is particularly evident for the lactate dehydrogenase (LDH) and the C-reactive protein (CRP), commonly associated with systemic inflammation. The increasing relationship between these markers and the WHO level indicates broader systemic involvement in cases of more severe COVID-19. Remarkably, pro-adrenomedullin (proADM) shows moderate variations across the WHO scale, despite being indicated as a good predictor of patient outcomes in previous studies [32, 33].

Finally, we observe that the age distribution does not show significant variations when stratifying patients using the WHO scale (Fig. 1-**d**). This is remarkable because it indicates that the risk of developing severe morbidity is not dependent on patients’ age. Additionally, we note that patients developing a mild form of COVID-19 typically arrive earlier at the hospital (have a lower Δ*t*_ons_) than patients experiencing a more severe illness progression (Fig. 1-**d**). We remark that the largest fraction of hospitalized patients reached level 3 on the WHO scale (see Fig. 1-**e**), which represents a critical threshold separating patients with mild to moderate symptoms from those with critical conditions. Importantly, this WHO level encompasses both patients with positive and negative outcomes, highlighting the challenge of accurately characterizing the severity of the disease based solely on clinical evidence.

### 2.2 The impact of age on the immune response

Aging is often characterized by immunosenescence, a phenomenon consisting of a decline in immune cell function and immune cell diversity, which reduces the capacity of patients to mount effective immune responses [34]. To uncover the signatures of immunosenescence in COVID-19 patients, we considered the rolling median of each feature over a 15-year half-window; the results are shown in Fig. 2. We also performed a quantitative comparison between the distributions of the features in the age range 40-70 years and 70-100 years, as 70 years approximately coincides with the median of the age distribution of inpatients (see Fig. 2-**d**).

**Figure 2:**
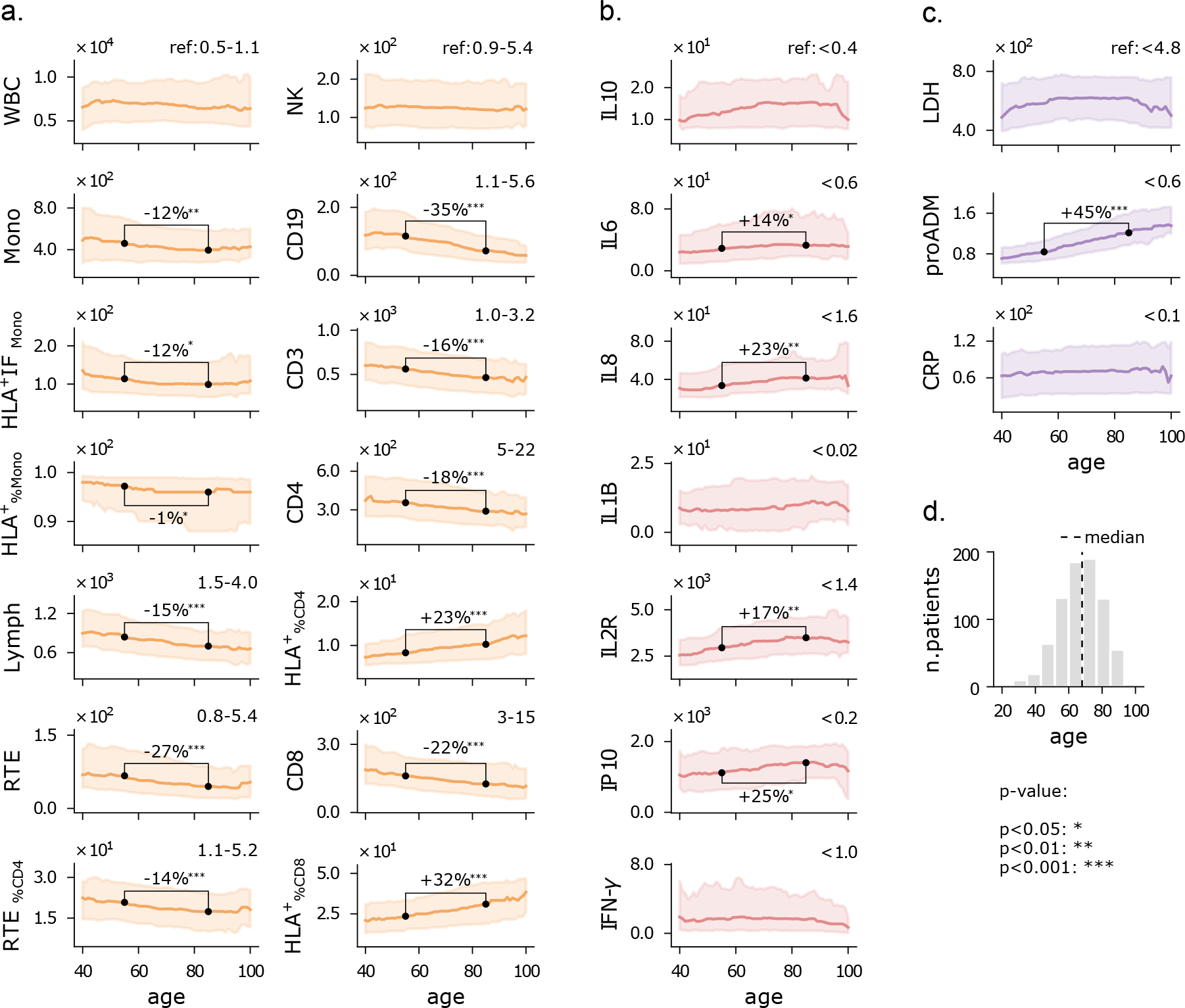
Features vs age. **a**) running median with a 15-year half-window of flow cytometry variables. The shaded area represents the first-to-third running quartiles. Two connected dots indicate the considered values to asses a (significant) change between age 55 and 85. **b**, **c**) similar to panel **a**, for cytokines and biomarkers, respectively. Reference values for healthy individuals are shown on the top-right of each plot, where available. **d**) age-histogram of the hospitalized patients. Measurement units as in Fig. 1.

We observe that the median of white blood cell (WBC) and natural killer cell (NK) counts remain constant across the age range at a value near the lower end of the reference range. Monocytes exhibit a slightly decreasing trend with age, both in terms of absolute number (Mono) and functionality (HLA^+^IF_Mono_). Lymphocytes show a marked decrease with age and are reduced compared to the reference value range, particularly recent thymic emigrants (RTE) and CD19 cells, implying that granulocytosis correlates with aging. Remarkably, the percentage of CD4 and CD8 cells expressing HLA-DR increases with age, indicating a larger decline in the number of inactive lymphocytes. Note that the average level of lymphocyte activity may still be reduced in older patients, and measurements of CD4 and CD8 intensity fluorescence (HLA^+^IF) would be required to confirm this observation. Moreover, the relative increase of HLA^+^_%CD4_ and HLA^+^_%CD8_ concomitant to a decrease in CD19 could reflect a cellular rather than humoral response. We point out that similar trends are observed for the younger cohort of outpatients, suggesting the presence of immunosenescence independently of the severity of the disease. However, outpatients present higher lymphocyte counts with a less pronounced decrease as they age, mainly pertaining RTE and CD19 cells, while their monocytes do not show significant age-related variations. Remarkably, they do not exhibit a reduction in CD4 and CD8 cells but still show an increase in HLA^+^_%CD4_ and HLA^+^_%CD8_ with age, like inpatients – see Fig. 7 in Appendix A. Overall, patients who did not necessitate hospitalization showed a stronger immune response and reduced variability with age compared to hospitalized patients.

Cytokine levels are consistently higher than reference values for all ages, which again reflects the dysregulated inflammatory state observed in COVID-19 patients. For most cytokines, there is a moderately increasing trend with age, particularly for IL8 and IL2R. This observation may stem from age-related low-grade chronic inflammation, which typically manifests as higher baseline cytokine levels [35].

All the inflammatory biomarkers exhibit median values exceeding the reference values, for patients of all ages. However, only the pro-adrenomedullin (proADM) displays a strong increasing relationship with age. Other studies have shown a significant interaction between proADM and age for clinical predictions, such as cardiovascular events [36], or more severe outcomes in COVID-19 patients [33], but the nature of this interaction remains an open question.

### 2.3 Temporal patterns of the immune response

The infection typically led COVID-19 patients to the hospital approximately 10 days after symptoms onset, with variations ranging from 1 day to more than 1 month. The heterogeneity in the time elapsed between symptoms onset and hospitalization, which we refer to as Δ*t*_ons_, reflects differences in the dynamics of disease progression. Small values of Δ*t*_ons_ indicate an acute onset that caused immediate hospitalization, while higher values suggest a slower disease where hospitalization is required only at a later stage.

To better understand the relationship between the immuno-inflammatory response and Δ*t*_ons_, we considered the rolling median of each feature with a 5-day half-window over the Δ*t*_ons_ dimension, thus providing a way to assess changes over time. Moreover, we compared the distributions of the features stratified within the ranges of 1-10 days and 11-20 days. We remark that our measurements do not come from the same patients over time, so they do not show the individual dynamics of the disease: the results displayed are a snapshot of the association between each variable and Δ*t*_ons_ and not an individual time series.

Despite the inherent noise associated with Δ*t*_ons_, an anamnestic variable, our analysis unveils clear patterns. Patients with high Δ*t*_ons_ show increased counts of white blood cells (WBC), monocytes, and CD19 cells, but reduced monocyte activation (HLA^+^IF_Mono_) and counts of natural killer cells (NK). In contrast, the number of T cells remains relatively constant across the entire Δ*t*_ons_ range. The observed increase in WBC counts is primarily driven by increased numbers of monocytes and granulocytes, likely compensating for the diminishing effectiveness of monocytes. Despite the low expression of natural killer and CD19 cells, we observed a standard trajectory of the immune response over time: a decrease in innate response activity (NK) paired with an increasing activation of the adaptive immune response system (CD19) as Δ*t*_ons_ increases.

The expression levels of cytokines broadly show a decreasing trend with Δ*t*_ons_, particularly IL10, IL1B, and IP10. The higher cytokine levels observed in patients with lower Δ*t*_ons_ suggest a more pronounced hyperinflammatory state, while lower values for larger Δ*t*_ons_ are compatible with a more subtle, but still higher-than-normal response. On the other hand, the observed dynamical pattern could be interpreted as a natural reduction of the inflammatory phase over time, and repeated measurements would be required to shed light on this phenomenon. Finally, the lactate dehydrogenase (LDH) trend indicates that patients arriving earlier at the hospital have a lesser degree of systemic damage. Similarly to pro-inflammatory cytokines, proADM tends to be higher in patients with lower Δ*t*_ons_.

Altogether, these results indicate that inflammation and cytokine levels are typically high when symptoms first appear, and they tend to be lower in patients developing a slower disease and arriving later at the hospital. This implies that patients hospitalized soon after the onset of symptoms typically exhibit a more acute condition, while those admitted later demonstrate advanced immune responses alongside more extensive tissue damage. Again, we shall remark that this is a cross-sectional analysis, and longitudinal data are needed for an accurate understanding of disease progression. Nonetheless, these observations highlight the importance of considering the immune response dynamics to assess patients’ clinical condition.

## 3 Outcome prediction analysis

The ability to mount an effective response to the infection strongly influences the prognosis of patients. In Sec. 2.1, we observed that reduced expression of lymphocytes and natural killer cells is typically associated with more severe disease progression, quantified in terms of the level reached on the WHO ordinal scale. However, the WHO scale only measures the morbidity level and does not explicitly carry information about patient outcomes such as mortality. Features identified as highly informative of disease severity may be less relevant for predicting patient outcomes and, thus, less useful for clinical decision-making. For instance, the amount of pro-adrenomedullin is weakly correlated with the severity level on the WHO scale (see Fig. 1-**e**), but it has been reported as a strong predictor for survival rate [11, 12]. To identify the immune-inflammatory markers with the highest predictive power for clinical outcome, we considered the problem of classifying patients based on the joint event of death and/or orotracheal intubation, designated as the ***death+OTI*** outcome; for a visual representation of the immuno-inflammatory features and clinical scores stratified by death+OTI, see Fig. 4. Note that this joint event has two advantages compared to mortality: it increases the number of records in the minority class (patients with negative outcome), and it mitigates the bias induced by clinical prioritization for invasive and life-saving interventions.

**Figure 3:**
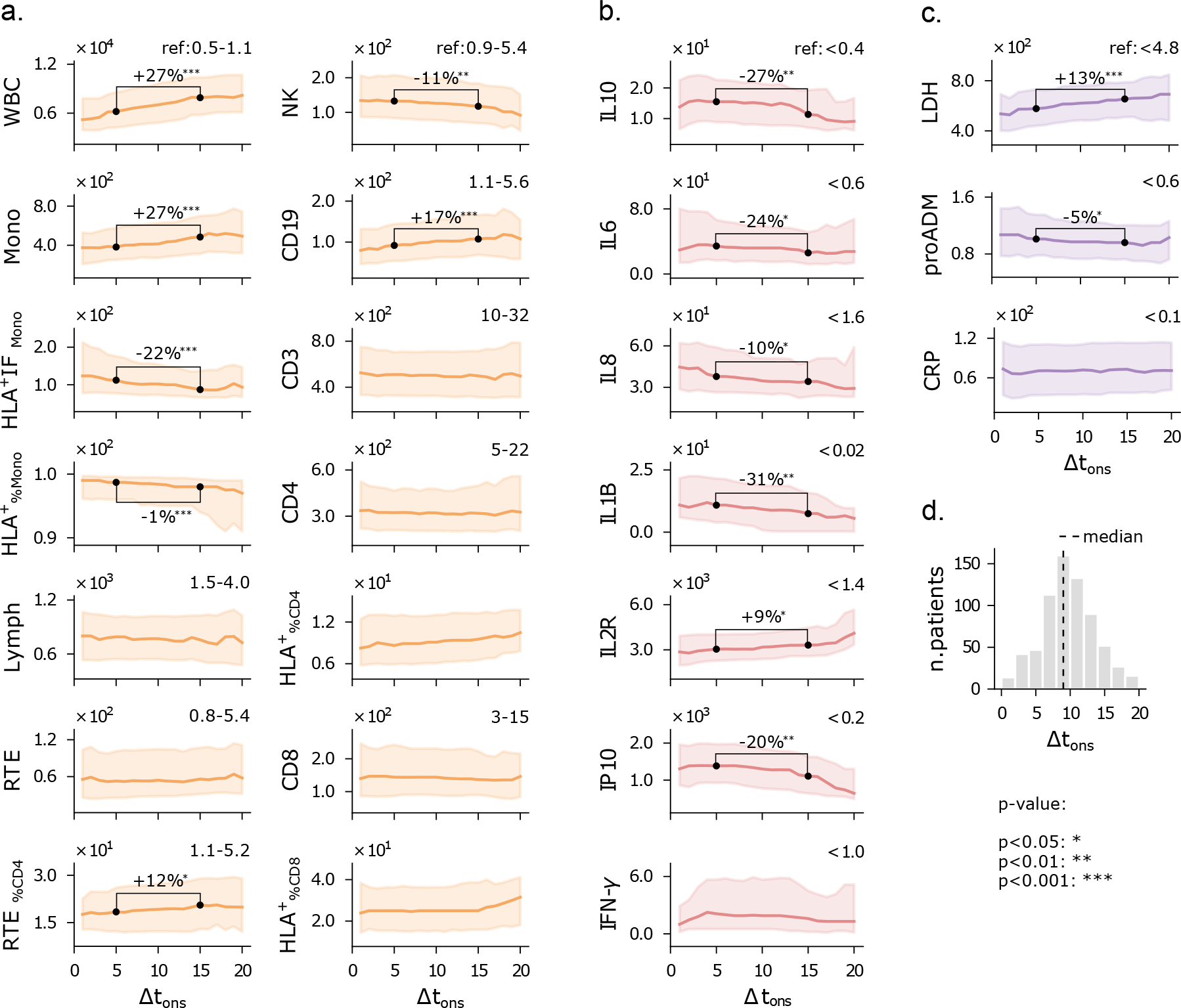
Features vs Δt_ons_. **a**) running median with a 5-day half-window of flow cytometry variables. The shaded area represents the first-to-third running quartiles. Two connected dots indicate the considered values to asses a (significant) change between Δ*t*_ons_ equal to 5 and 15 days. **b**, **c**) similar to panel **a**, for cytokines and biomarkers, respectively. Reference values for healthy individuals are shown on the top-right of each plot, where available. **d**) Δ*t*_ons_-histogram of the hospitalized patients. Measurement units as in Fig. 1.

**Figure 4:**
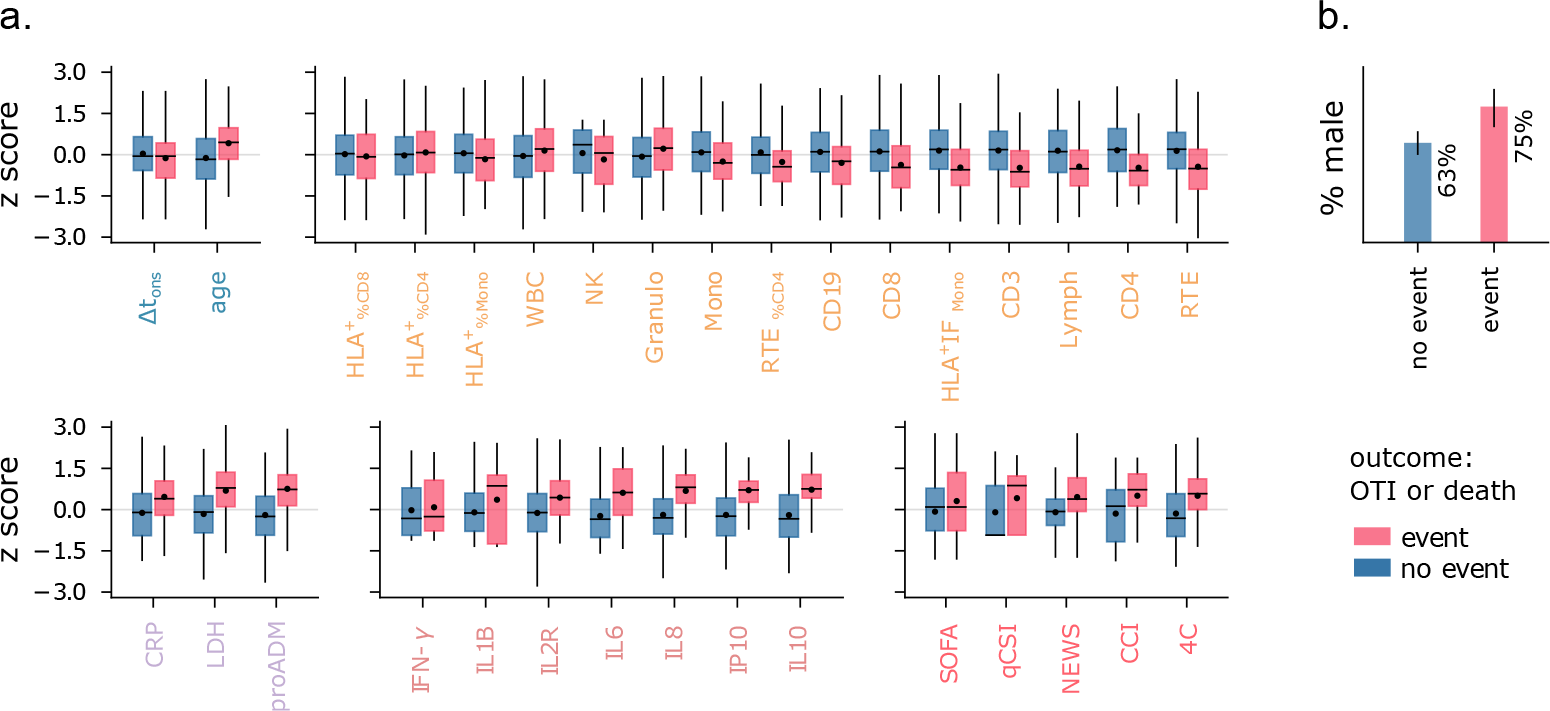
Features vs outcome. **a**) Distribution of normalized features stratified by outcome. Boxes include first to third quartiles. The horizontal bar and black dot indicate the median and mean of the distribution, respectively. **b**) Percentage of male patients with a positive (blue) and negative (red) outcome.

We based our analysis on logistic regression (LR) models, as preliminary results with non-linear classifiers such as support vector machines and random forests did not show significant improvements. We conducted two types of assessments of the LR models: the overall predictive performance, measured by the area under the ROC curve (AUC), and the ability to identify low-risk patients. Specifically, we considered the task of detecting the largest number of patients who did not experience the event while maintaining the fraction of false negatives below a predetermined low value. This second assessment is motivated by the necessity to establish priority rules for clinical and hospital operations, which become crucial during critical situations such as a pandemic outbreak. We set a minimum negative predictive value (NPV) requirement of 0.97 for the classifiers on the training set, and we considered NPV values above 0.95 acceptable for the test set. These thresholds are illustrative, but it is important to note that NPV during testing is typically lower than the value set as a requirement during training. Hence, the latter needs to be adjusted to account for this discrepancy. Detailed information on data preprocessing and model specifications can be found in Sec. 5.

### 3.1 Cytokine-based univariate models outperform baseline indices

Firstly, we evaluated the predictive power of individual features using univariate LR models, removing missing data from the analysis. The results are displayed in Fig. 5. The Charlson Comorbidity Index (CCI) and the 4C score show the highest AUC among the clinical indices. In the following, we will refer to the 4C score as the best baseline predictor and use the associated AUC as a reference value (AUC_ref_ = 0.69). Remarkably, the flow cytometry features, cytokines, and biomarkers all include variables that match or outperform the best baseline. Lymphocytes (RTE, CD3, CD4, Lymph) and monocyte activation (HLA^+^ IF_Mono_) are the strongest predictors among the flow cytometry features, with the AUC of RTE and CD3 matching AUC_ref_. We observe that granulocytes tend to be more expressed in patients with a negative outcome (see Fig. 4), contrary to monocytes and lymphocytes. For this reason, the absolute count of white blood cells has low predictive power compared to these three WBC subpopulations. It is important to note that we did not measure granulocyte counts directly; instead, we estimated them by subtracting lymphocyte and monocyte counts from the total white blood cell count. The AUC_ref_ value is significantly outmatched by the mid-regional pro-adrenomedullin (proADM), which is confirmed as a strong predictor of clinical outcome, and by several cytokine expression levels. In particular, IL8 and IL10 have the highest AUC scores of all the univariate LR models (AUC = 0.79). IL8 is a potent, proinflammatory chemokine that induces degranulation of neutrophils and adhesion of polymorphonuclear cells to the endothelium, and it is released from several cell types in response to inflammation, including monocytes, macrophages and neutrophils [37]. This pro-inflammatory mechanism mediated by IL8 may underlie the joint increase in granulocytes and IL8 concentrations observed in patients with poor outcomes. Instead, IL10 plays an essential role in inducing an immunoregulatory phenotype in B cells that exerts substantial anti-inflammatory and immunosuppressive functions [38].

**Figure 5:**
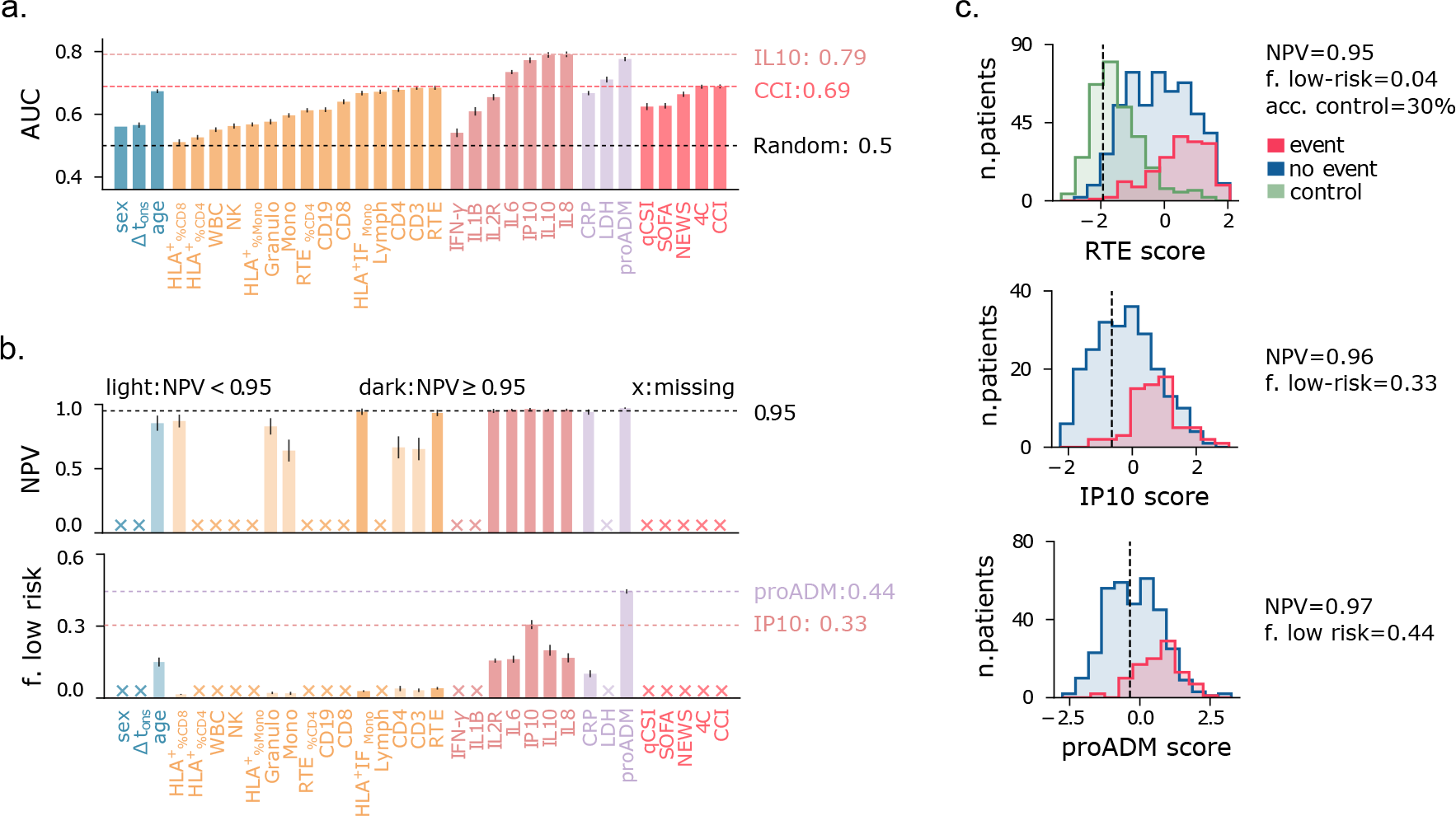
Univariate models. **a**) Area under the ROC curve (AUC) of univariate logistic regression (LR) models predicting the event death+OTI. Input variables are shown on the x-axis. Error bars show the 95% confidence intervals. **b**) Negative predictive value (NPV) and fraction of low-risk patients correctly identified (specificity), conditional on NPV *≥* 0.97 on the training set. Crosses on the x-axis indicate models not satisfying this requirement. Light-colored bars indicate models with test NPV below 0.95. **c**) LR score histograms of patients with positive (blue) and negative (red) outcomes, and of outpatients (green, control), for the best predictors among flow cytometry features, cytokines, and biomarkers. The black vertical line marks the cutoff threshold for the minimum NPV requirement.

Finally, we confirm that age is a relatively good predictor, as often reported [39, 40], with an AUC slightly below AUC_ref_. Sex and Δ*t*_ons_, instead, have predictive power significantly below the best baseline predictor. To summarize, a negative outcome is strongly associated with high cytokine expression, increased endothelial dysfunction (high proADM), as well as cell injury (high LDH), and immune deficiency (IL10 overexpression, reduced lymphocytes, monocytes, and monocyte activation).

Identifying low-risk patients with high confidence using only one feature at a time poses a significant challenge, and none of the baseline clinical indices proves useful for this task (see Fig. 5-**b**). Instead, the proADM-based model achieves a remarkable result, identifying 44% of negative cases with a false omission rate of only 3%. Low-risk patients can also be identified using cytokine expression levels, particularly IP10 (specificity = 0.31). Instead, among all flow cytometry features, only the recent thymic emigrants (RTE) and monocyte activation (HLA^+^ IF_Mono_) features fulfill the minimum NPV requirement on the test set, but with low specificity. The identified low-risk cutoff values are reported in Table 2. The score histogram of RTE, IP10, and proADM-based models are depicted in Fig. 5-**c**, where the vertical dashed line marks the threshold below which patients are considered at low risk. Remarkably, only 30% of outpatients would be correctly classified as low-risk with the RTE-based predictor. As we will see, this fraction improves dramatically when using multivariate models.

**Table 2:** Thresholds for low-risk patient identification. Observed range (1^st^ *−* 99^th^ quantiles), low-risk cutoff threshold, and associated negative predictive value (NPV) and specificity of univariate predictors. The best low-risk predictor (proADM) is highlighted in bold. Measurement units: RTE: U*/µ*L, cytokines: pg*/*mL; proADM: nmol*/*L; CRP: mg*/*L.

| | RTE | IL10 | IL2R ($10^3$) | IL6 | IL8 | IP10 ($10^2$) | proADM | CRP |
| --- | --- | --- | --- | --- | --- | --- | --- | --- |
| **range** | 3 - 346 | 2.3 - 85.7 | 0.8 - 11.0 | 2 - 504 | 9 - 255 | 1 - 36 | 0.45 - 3.34 | 1 - 269 |
| **cutoff** | $\geq 263$ | $\leq 5.5$ | $\leq 1.9$ | $\leq 6.9$ | $\leq 19$ | $\leq 6.5$ | **$\leq 0.84$** | $\leq 9$ |
| **NPV** | 0.93 | 0.96 | 0.95 | 0.96 | 0.96 | 0.96 | 0.97 | 0.94 |
| **specif city** | 0.04 | 0.20 | 0.16 | 0.16 | 0.17 | 0.31 | **0.44** | 0.10 |

### 3.2 Multivariate models improve identification of low-risk patients

The immuno-inflammatory features provide complementary information on the host’s response to the infection and, when combined, can lead to more accurate predictions. To this end, we employed multivariate LR models considering the sets of features FC=(CD3, RTE %_CD4_, CD19, Mono, HLA^+^FI _Mono_, Granulo), CK=(IL1B, IL2R, IL6, IL8, IL10), and BM=(CRP, LDH, proADM). We used these sets individually, or jointly with the set Dem=(sex, age, Δ*t*_ons_). Note that FC and CK are subsets of the sets introduced in Sec. 2 with the same names; these variables were chosen for their clinical relevance, while also maintaining predictive power and controlling for collinearities. Only cases with less than 50% of missing data for each considered set of variables were included in the multivariate analysis. The resulting datasets, described in Supp. Table 3, had missing values replaced via KNN imputation.

The results of the multivariate prediction are summarised in Fig. 6, where panel **a** shows the AUC of multivariate logistic regression models, and of the univariate models employing variables in the FC, CK, BM, and Dem sets. The best performances are again obtained using cytokines and serological biomarkers. In particular, the CK+Dem model has AUC = 0.87, a large improvement over the best baselines (AUC_ref_ = 0.69), and it shows the best performance in identifying low-risk patients, with specificity = 0.59 and NPV = 0.96 (panel **b**). This is remarkable, as it means that we could correctly detect 59% of low-risk patients with only 4% of false negative calls. The FC+Dem (AUC = 0.78) and BM+Dem (AUC = 0.81) models also show significant improvements over the best baselines. Fig. 6-**c** provides a visual representation of these results with score histograms for the FC+Dem, BM+Dem, and CK+Dem predictors. Note that, for the FC+Dem model, the threshold for low-risk patients gives an accuracy of 86% on the control set. This is a striking improvement over the best univariate predictor from the FC set, the RTE-based model, which accurately classifies only 30% of control patients when using the same criterion (Fig. 5-**c**). The relative importance of each feature in the FC+Dem score is depicted in Fig. 6-**d**, which shows the normalized coefficients of the model. Large positive (resp. negative) values imply a strong positive (resp. negative) association with the outcome. Note that all variables are important for the score, as all coefficients differ from zero. The most important features negatively associated with the outcome are HLA^+^ IF_Mono_, and CD3, while positive associations with the outcome are observed for age and granulocytes. Ranking variables by weight magnitude does not give a one-to-one match with univariate AUC ranks (Fig. 5-**a**). For example, monocyte activation (HLA^+^ IF_Mono_) is the 4^th^ best univariate predictor, while it represents the most important variable of the FC set. This indicates that the FC+Dem model is able to capture non-trivial interactions among the features. From this picture, it is clear that the state of cells involved in the immune response, measured upon hospitalization, can be highly informative of disease progression. However, we shall remark that the most significant feature in the FC+Dem model is age, a proxy variable that does not directly relate to the host’s response. This is not the case for the BM+Dem and CK+Dem models (Fig. 6-**e**, **f**), validating the observation that cytokines and inflammatory biomarkers hold the most relevant information about the disease progression. Finally, we observe that sex and Δ*t*_ons_ show negative associations with the outcome across all models (Fig. 6-**d**-**f**), suggesting a higher likelihood of a positive outcome for females, as frequently noted [41, 42], and for patients hospitalized later after symptoms onset.

**Figure 6:**
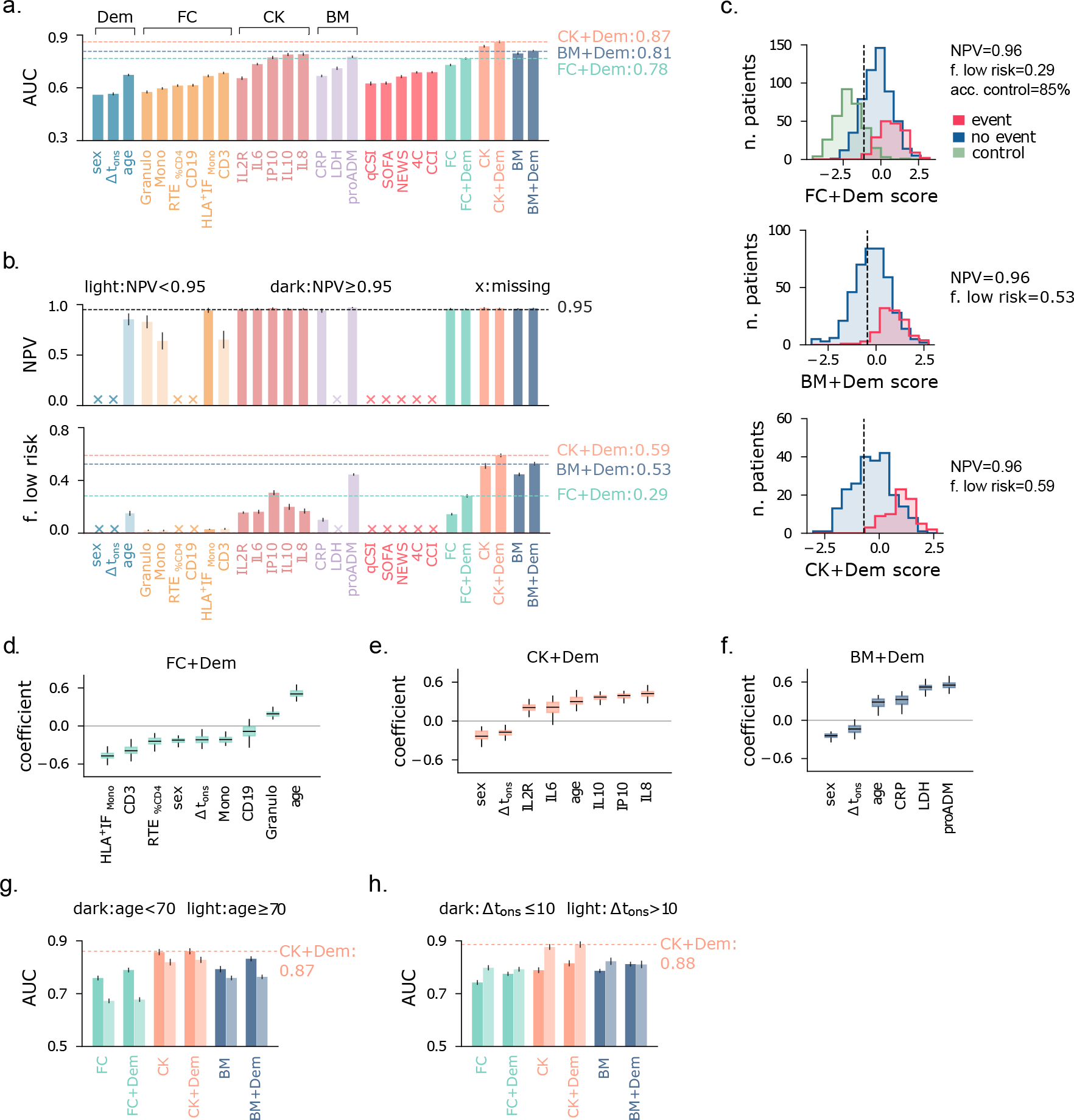
Multivariate models. **a**) Area under the ROC curve (AUC) of univariate and multivariate logistic regression (MLR) models predicting the event death+OTI. MLR models are named according to the set of input variables. Error bars show the 95% confidence intervals. **b**) Negative predictive value (NPV) and fraction of low-risk patients correctly identified (specificity), conditional on training NPV *≥* 0.97. Crosses indicate models not satisfying this requirement. Light-colored bars indicate models with test NPV below 0.95. **c**) MLR score histograms of inpatients with positive (blue) and negative (red) outcomes, and of outpatients (green, control). The vertical line marks the cutoff threshold for the minimum NPV requirement. **d**–**f**) Normalized coefficients box plots of main MLRs. **g**, **h**) AUC of MLRs stratified by age and Δ*t*_ons_.

### Outcome prediction across population strata

We conclude our analysis by examining the results of outcome prediction upon stratifying the population by age (≶ 70 years) and Δ*t*_ons_ (≶ 10 days). First of all, we note that predicting the outcome becomes harder for patients aged above 70 years, as individuals in this age range are more fragile. Indeed, models using flow cytometry features (FC, FC+Dem) and biomarkers (BM, BM+Dem) have significantly higher AUC for the younger cohort (Fig. 6-**g**). The multivariate predictors based on cytokines (CK, CK+Dem) show a less marked difference in AUC across the two age strata, reaching AUC = 0.87 for the younger cohort. When stratifying patients by Δ*t*_ons_, predicting the outcome is easier for patients with Δ*t*_ons_ *>* 10, likely due to their more advanced response, which provides clearer prognostic indications. In particular, the cytokines-based models exhibit the largest variations in AUC across strata, with a remarkable AUC = 0.88 for patients with Δ*t*_ons_ *>* 10 (Fig. 6-**h**). Therefore, for patients hospitalized later after symptoms onset, cytokine levels can be highly informative of outcome severity. As expected, including features from the Dem set only yields minor improvements upon stratification over age and Δ*t*_ons_.

## 4 Discussion

This paper provides a comprehensive description of the immuno-inflammatory response observed in COVID-19 patients before vaccination, thus serving as a case study for a first-time infection by a new virus. Our analysis integrates white blood cell subpopulations, cytokine expression levels, and serological biomarkers measured at hospital admission, offering a multifaceted description of several components of the immune system and inflammatory pathways. Analyzing these features together constitutes one of the core strengths of our contribution, as they are typically studied separately in the literature. From an overall standpoint, we observed that lymphopenia, compromised monocyte function, granulocytosis, and heightened cytokine levels correlated with disease severity and a negative outcome. Older patients exhibited a more compromised immune response characterized by reductions in monocytes, T cells, and B cells, suggestive of immunosenescence. Remarkably, they also showed increased percentages of HLA-DR-positive T cells. Similar patterns were observed in the cohort of asymptomatic outpatients, which, however, maintained higher lymphocyte levels, with only recent thymic emigrants and B cells decreasing with age. Additionally, cytokine expression levels showed a weak positive correlation with age, while mid-regional pro-adrenomedullin (proADM) levels strongly correlated with age, possibly due to underlying chronic low-grade inflammation and endothelial dysfunction that develops with aging. To investigate the temporal behavior of the immune response, we considered the number of days elapsed between symptom onset and hospitalization (Δ*t*_ons_). Our cross-sectional analysis revealed a decrease in the effectiveness of monocytes over time, characterized by an increase in their numbers but a concurrent decrease in their activation level, as measured by the mean intensity fluorescence of HLA-DR. We also observed a reduction in natural killer cells and cytokine levels, higher concentrations of lactate dehydrogenase (LDH), a marker of cell injury, and increased activation of the humoral response, as indicated by B CD19 cells. This pattern mirrors a typical dynamic evolution of the host’s immune response, suggesting that these results should be interpreted longitudinally. On the other hand, patients with different Δ*t*_ons_ may manifest intrinsically different diseases, where some exhibit a rapid and intense response and others undergo a slower but more subtle progression, leading to the need for hospital care at a later stage.

The serum levels of cytokines, whether proinflammatory (IL8, IP10) or immunomodulatory (IL10), emerged as the most crucial aspect of the immuno-inflammatory response for predicting patients’ outcomes. Indeed, cytokine-based logistic regression models substantially outperformed the best baseline index, the 4C score, developed to predict in-hospital mortality for COVID-19 patients. Additionally, pro-adrenomedullin proved to be the best individual predictor for identifying low-risk patients, with a cutoff of *≤* 0.84 nmol*/*L corresponding to a specificity of 0.44 and a negative predictive value (NPV) of 0.97. Importantly, this cutoff aligns well with those found in other settings [11, 43], underscoring the robustness of this result. However, we shall remark that proADM is weakly correlated with the severity level on the WHO scale, and it may reflect a broader physiological response, not necessarily specific to the severity of COVID-19. Therefore, using proADM alone for risk stratification might not fully address treatment needs specific to COVID-19, such as oxygen-based therapies. In contrast, cytokine expression levels may provide a more targeted tool for identifying low-risk patients, as they show significant correlations with both COVID-19 morbidity and mortality. Notably, when combining cytokine levels with demographic information into a single model, we achieved a remarkable AUC of 0.87. This model also proved very effective in detecting low-risk patients, reliably identifying approximately 60% of individuals who may not require hospitalization, while maintaining a false omission rate of just 4%. Moreover, our analysis revealed that cytokine-based models can be further improved when focusing on patients hospitalized later after symptom onset (Δ*t*_ons_ *>* 10). This finding, if interpreted longitudinally, suggests that monitoring the evolution of the cytokine levels through follow-up tests could offer valuable predictive insights. Lastly, our analysis shows that flow cytometry features exhibit less predictive power than cytokines (IL6, IP10, IL10, IL8) and biomarkers (LDH, proADM). Nevertheless, features such as recent thymic emigrants and T CD3 cells match the performance of the best baseline predictor, the 4C score. When integrated into a single model, the flow cytometry features outperform the 4C score, leveraging non-trivial relationships between white blood cell subpopulations and thus underscoring the complexity of the immune response. Notably, in the combined model, the mean intensity fluorescence (IF) of monocytes HLA-DR emerges as the most important flow cytometry feature, highlighting the critical role of cell activation in the immune response. Exploring HLA-DR IF in lymphocyte subpopulations could further improve our understanding of the immune response to COVID-19 and other infectious diseases.

### 4.1 Limitations

Despite the valuable insights and contributions provided by this study, several limitations need to be acknowledged. Firstly, our dataset included measurements of cytokine expression levels and serological biomarkers only for subsets of hospitalized patients, with no corresponding measurements for outpatients or healthy individuals. Therefore, our control set only encompassed demographic information and flow cytometry measurements, and we had to rely on reference values for cytokine expressions and inflammatory biomarkers. Moreover, due to the limited availability of information regarding cytokines and biomarkers, we opted to employ multivariate logistic regression models where these features are not combined. This approach aimed to prevent further reduction in dataset size and maintain statistical power in our analyses. Combining all covariates into a single multivariate predictor could potentially improve predictive power, though it would require more advanced feature selection methods to enhance model interpretability [44]. Another limitation is the absence of external validation for our predictive models. While our study utilized a large dataset from our hospital, the generalizability of our findings to other healthcare settings, populations, and other infectious diseases remains uncertain. As already reported in the text, it is important to acknowledge the limitation of the Δ*t*_ons_ variable, which relies on the ability of patients to accurately recall when symptoms onset occurred. This variable is thus subject to significant variability stemming from differences in symptom perception and memory capabilities of the patients. Furthermore, we remark that our study was conducted within a specific context and timeframe, involving patients admitted to our hospital during the COVID-19 pandemic before the vaccination campaign started. As the understanding of COVID-19 and its management continues to evolve, future studies incorporating diverse populations and settings are needed to validate and expand upon our findings. Finally, like any observational study, our research is subject to potential confounders and unmeasured variables. Despite our best efforts to account and adjust for known factors, there may still be uncontrolled variables that could influence the results.

## 5 Materials and methods

### Data collection

This study involves data from patients admitted to the Infectious Disease ward of the Azienda Sanitaria Universitaria Friuli Centrale Santa Maria della Misericordia of Udine, a 1000-bed tertiary-care teaching hospital identified as a regional referral center for COVID-19 patients. The analyzed records were collected from March 2020 to April 2021, covering the first, second, and part of the third pandemic waves. During this period, clinical data from patients admitted for SARS-CoV-2 were included in a retrospective registry, namely the “MAnagement coroNavirus Disease In hospital (MANDI) registry” (authorization of DG, decree n. 957, 10/09/2021). Patients were enrolled in accordance with the Helsinki Declaration. Ethical approval was granted from governance bodies of Friuli Venezia Giulia. The registry included patients admitted to either the infectious disease clinic or the intensive care unit, diagnosed with SARS-CoV2 infection through at least one positive nasopharyngeal swab, confirmed by reverse transcriptase PCR assays. The eligible participants were aged 18 years or older and had provided informed consent for the utilization of anonymous clinical data. Hospital admission involved routine inquiries regarding consent for anonymized aggregate data for research purposes, facilitated by the General Electronic Consents (GECO system). Relevant patients’ data were extracted by a team of physicians from the hospital electronic health record (INSIEL, Trieste, Italy), anonymized, and recorded on a cloud-based clinical data management platform (Castor, Netherlands and USA). ***All patients had not yet been vaccinated against SARS-CoV-2***.

### Immuno-inflammatory features

Approximately 900 patients underwent lymphocyte and monocyte immunophenotyping within the first 72 hours of hospital admission. The following measurements were collected via multicolor flow cytometry: absolute number of white blood cells (WBC), lymphocytes (Lymph), monocytes (Mono), T CD3^+^ cells (CD3), Th CD3^+^CD4^+^ cells (CD4), Tc CD3^+^CD8^+^ cells (CD8), T NK CD3^−^CD56/CD16^+^ cells (NK), B CD3^−^CD19^+^ cells (CD19), and recent thymic emigrants (RTE), as well as the percentage of HLA-DR-positive monocytes (HLA^+^_%Mono_), CD4 (HLA^+^_%CD4_), and CD8 (HLA^+^_%CD8_) cells, the percentage of RTE-CD4 cells (RTE _%CD4_), and the monocytes HLA-DR mean intensity fluorescence (HLA^+^IF _Mono_). We refer to the flow cytometry features as the ***FC set***. The same measurements were taken for approximately 370 asymptomatic outpatients and 90 healthy individuals. These two cohorts exhibit matching statistics (see Supp. Fig. 8), and we employed the outpatients’ data as a control dataset for our analysis. The flow cytometry features provide a comprehensive description of the immune response, encompassing the innate immune system (Mono, NK), the cell-mediated adaptive system (CD4, CD8), and the antibody-mediated adaptive system (CD19). We remark that many of these features are hierarchically related: WBCs consist of lymphocytes, monocytes, and granulocytes. Lymphocytes include CD3, CD19, and NK cells. CD3 cells can be categorized into CD4 and CD8 cells, and RTE cells are a subpopulation of CD4 cells. It is important to consider these relationships to avoid collinearity effects.

In our analysis, we also employed cytokine expression levels and serological inflammatory biomarkers. The measured cytokines included interleukin 10 (IL10), IL1-Beta (IL1B), sIL2R-*α*/sCD25 (IL2R), interleukin 6 (IL6), interleukin 8 (IL8), chemokine IP10/CXCL10 (IP10), and interferon-*γ* (INF-*γ*). Note that this set of features, named the ***CK set***, encompass both pro-inflammatory signals (IL6, IL8, IP10) and immunomodulatory ones (IL10, IL2R) [14]. Cytokines were analyzed using a microfluidic ultrasensitive ELISA assay with the Protein Simple Plex technology on the Ella Instrument (R&D systems, Bio-Techne, USA). Regarding the serological biomarkers, we measured pro-adrenomedullin (proADM), lactate dehydrogenase (LDH), and C-reactive protein (CRP). These features are referred to as the ***BM set*** and provide information about the host’s inflammatory state. More specifically, LDH is a marker of cell injury that can be employed to assess lung damage [45], proADM is a marker of endothelial damage and a good predictor of COVID-19 severity [11], and CRP, a standard inflammatory indicator, may signal a severe systemic response to the infection.

In addition to the immuno-inflammatory features described above, we collected information on several comorbidities for all patients enrolled in the study, as well as age, sex, and number of days elapsed between symptoms onset and hospitalization. We refer to Supp. Table 4 for a detailed description of the dataset.

### Preprocessing and statistical analysis

Manually filled databases often contain typos, leading to the presence of outliers or attributes with no physical meaning. To minimize the impact of these mistakes and ensure the data’s homogeneity and cleanliness, we implemented the following pre-processing steps: we considered patients with age in the range of 30 to 100 years and Δ*t*_ons_ between 0 and 30 days. We further filtered records to include only patients with a CCI score of less than 7, to mitigate potential confounding factors for the outcome of the patients and ensure a more accurate description of the immune response. High CCI scores indicate the presence of significant comorbidities, which could interfere with the interpretation of immunological data and may lead to an inaccurate representation of the true effects of COVID-19 on the immune system. Finally, we performed a careful inspection of the distributions of features and a conservative outlier removal step. To detect outliers, we employed a power transform from scikit-learn [46] to normalize the data distribution. We then set to NAN data with a z-score of absolute value larger than 3.

Our characterization of the immuno-inflammatory profiles entailed multi-dimensional comparisons among various patient groups. More precisely, we investigated the relationship between features and developed disease severity by stratifying patients according to the ***WHO scale*** (Fig. 1). This scale is based on the World Health Organization’s guidance [16] and categorizes patients into four levels as follows:

**1. *mild disease*** : symptomatic patients without pneumonia;
**2. *moderate disease***: patients with clinical signs of pneumonia with no need for oxygen therapy;
**3. *severe disease***: patients with clinical signs of severe pneumonia in need of oxygen therapy;
**4. *critical disease*** : patients with oxygenation impairment, acute respiratory syndrome, and/or sepsis.

To detect relevant patterns of the immune response, we also analyzed the rolling median of all features along two axes: the age and Δ*t*_ons_ of the patients. More precisely, we employed a half-window of 15 years to examine age-related variations (Fig. 2), comparing the median of two age groups: patients aged 40-70 and 70-100 years. Similarly, we used a half-window of 5 days for the Δ*t*_ons_ (Fig. 3) and compared patients with Δ*t*_ons_ of 1-10 and 11-20 days. The distributions of different population groups were tested with the Mann-Whitney U test.

### Outcome prediction

In Sec. 3, we explored the potential of predicting patient outcomes based on their immune response as measured upon hospital admission. To this end, we employed both univariate and multivariate logistic regression (LR) models to predict the joint event of death and/or orotracheal intubation, referred to as the ***death+OTI*** outcome. Combining these two events helps to mitigate an important source of bias: patients’ prioritization for invasive and life-saving interventions. Moreover, considering this outcome offers a second advantage: it improves class imbalance, with a negative event observed in approximately 24% of patients, compared to 17% when considering death only (Supp. Table 4). To address the class imbalance, we used logistic regression models from scikit-learn with *l*_2_ penalty and class weights balanced based on class frequency. We evaluated univariate LR models by removing records with missing data, thus capturing the true signal associated with each variable. We randomly split records into train (70%) and test (30%) sets, keeping the class and sex frequencies unchanged, and repeating the process several times to collect performance statistics. For the multivariate models, we considered different subsets of features, controlling for collinearity via the variance inflation factor and domain-based knowledge. Records with more than 50% of missing data in each feature subset were omitted from the analysis. The remaining missing data were replaced via KNN imputation (*K* = 10, neighbors weighted uniformly), and the least significant PCA components (with less than 5% of explained variance) were removed from the data. We combined k-fold cross-validation and grid-search on each train set to select the regularization strength for the *l*_2_ penalty.

We measured the performance of each logistic regression model in two ways. First, we computed the area under the ROC curve (AUC) to quantify the overall predictive power of the classifier. Second, we evaluated the ability of each classifier to detect the largest fraction of low-risk patients, i.e., patients with a positive outcome, while retaining a low false omission rate. Practically, we set a requirement of a negative predictive value (NPV) of 0.97 on the train set, with NPV values above 0.95 considered acceptable on the test set, and we measured the performance in detecting low-risk patients in terms of specificity.

## Reproducibility

The code and dataset to reproduce our results are available here.

## Data Availability

All data produced are available online on a public GitHub repository.

https://github.com/RiccardoGM/ImmuneResponseCOVID19

## A Supplementary material

**Table 3:**
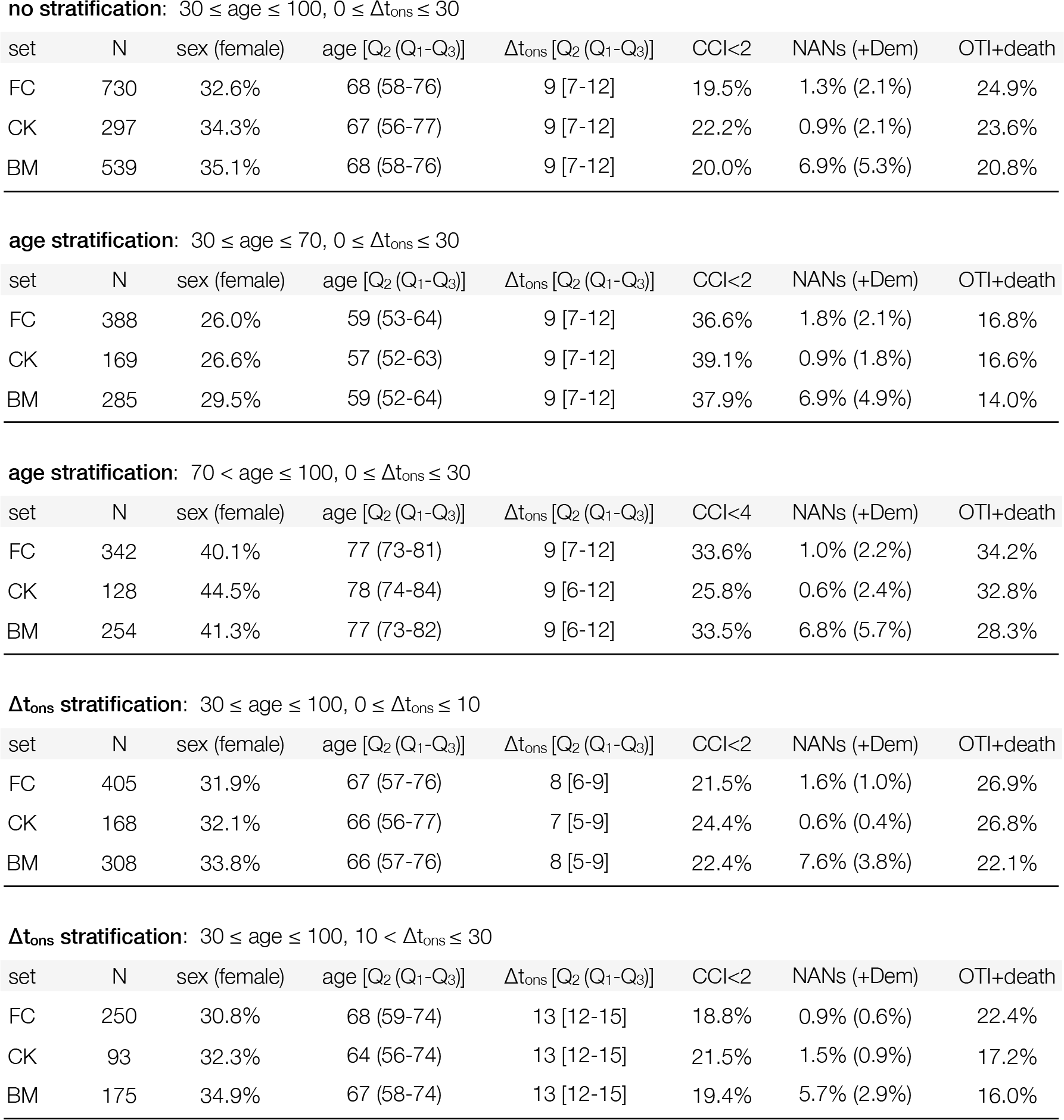
Datasets employed for the multivariate predictions. Demographics and clinical characteristics of the datasets utilized in the multivariate predictive analysis described in Sec. 3.2. Records are removed if more than 50% of data is missing in any feature set included among the predictors. The considered feature sets incorporate flow cytometry variables (FC set), cytokines (CK set), serological inflammatory biomarkers (BM set), and demographic information (Dem set). More than 50% of the demographic data is available for all records, so combining the Dem set with any on the FC, CK, or BM set does not decrease the size of the considered dataset. Stratified datasets do not include records with missing information in the stratification variable.

| set | N | sex (female) | age [Q<sub>2</sub> (Q<sub>1</sub>-Q<sub>3</sub>)] | $\Delta t_{\text{ons}}$ [Q<sub>2</sub> (Q<sub>1</sub>-Q<sub>3</sub>)] | CCI<2 | NANs (+Dem) | OTI+death |
| --- | --- | --- | --- | --- | --- | --- | --- |
| FC | 730 | 32.6% | 68 (58-76) | 9 [7-12] | 19.5% | 1.3% (2.1%) | 24.9% |
| CK | 297 | 34.3% | 67 (56-77) | 9 [7-12] | 22.2% | 0.9% (2.1%) | 23.6% |
| BM | 539 | 35.1% | 68 (58-76) | 9 [7-12] | 20.0% | 6.9% (5.3%) | 20.8% |

**age stratification:** $30 \leq \text{age} \leq 70$ , $0 \leq \Delta t_{\text{ons}} \leq 30$
| set | N | sex (female) | age [Q<sub>2</sub> (Q<sub>1</sub>-Q<sub>3</sub>)] | $\Delta t_{\text{ons}}$ [Q<sub>2</sub> (Q<sub>1</sub>-Q<sub>3</sub>)] | CCI<2 | NANs (+Dem) | OTI+death |
| --- | --- | --- | --- | --- | --- | --- | --- |
| FC | 388 | 26.0% | 59 (53-64) | 9 [7-12] | 36.6% | 1.8% (2.1%) | 16.8% |
| CK | 169 | 26.6% | 57 (52-63) | 9 [7-12] | 39.1% | 0.9% (1.8%) | 16.6% |
| BM | 285 | 29.5% | 59 (52-64) | 9 [7-12] | 37.9% | 6.9% (4.9%) | 14.0% |

**age stratification:** $70 < \text{age} \leq 100$ , $0 \leq \Delta t_{\text{ons}} \leq 30$
| set | N | sex (female) | age [Q<sub>2</sub> (Q<sub>1</sub>-Q<sub>3</sub>)] | $\Delta t_{\text{ons}}$ [Q<sub>2</sub> (Q<sub>1</sub>-Q<sub>3</sub>)] | CCI<4 | NANs (+Dem) | OTI+death |
| --- | --- | --- | --- | --- | --- | --- | --- |
| FC | 342 | 40.1% | 77 (73-81) | 9 [7-12] | 33.6% | 1.0% (2.2%) | 34.2% |
| CK | 128 | 44.5% | 78 (74-84) | 9 [6-12] | 25.8% | 0.6% (2.4%) | 32.8% |
| BM | 254 | 41.3% | 77 (73-82) | 9 [6-12] | 33.5% | 6.8% (5.7%) | 28.3% |

**$\Delta t_{\text{ons}}$ stratification:** $30 \leq \text{age} \leq 100$ , $0 \leq \Delta t_{\text{ons}} \leq 10$
| set | N | sex (female) | age [Q<sub>2</sub> (Q<sub>1</sub>-Q<sub>3</sub>)] | $\Delta t_{\text{ons}}$ [Q<sub>2</sub> (Q<sub>1</sub>-Q<sub>3</sub>)] | CCI<2 | NANs (+Dem) | OTI+death |
| --- | --- | --- | --- | --- | --- | --- | --- |
| FC | 405 | 31.9% | 67 (57-76) | 8 [6-9] | 21.5% | 1.6% (1.0%) | 26.9% |
| CK | 168 | 32.1% | 66 (56-77) | 7 [5-9] | 24.4% | 0.6% (0.4%) | 26.8% |
| BM | 308 | 33.8% | 66 (57-76) | 8 [5-9] | 22.4% | 7.6% (3.8%) | 22.1% |

Table 3: ***Datasets employed for the multivariate predictions.*** Demographics and clinical characteristics of the datasets utilized in the multivariate predictive analysis described in Sec.
| set | N | sex (female) | age [Q<sub>2</sub> (Q<sub>1</sub>-Q<sub>3</sub>)] | $\Delta t_{\text{ons}}$ [Q<sub>2</sub> (Q<sub>1</sub>-Q<sub>3</sub>)] | CCI<2 | NANs (+Dem) | OTI+death |
| --- | --- | --- | --- | --- | --- | --- | --- |
| FC | 250 | 30.8% | 68 (59-74) | 13 [12-15] | 18.8% | 0.9% (0.6%) | 22.4% |
| CK | 93 | 32.3% | 64 (56-74) | 13 [12-15] | 21.5% | 1.5% (0.9%) | 17.2% |
| BM | 175 | 34.9% | 67 (58-74) | 13 [12-15] | 19.4% | 5.7% (2.9%) | 16.0% |

**Figure 7:**
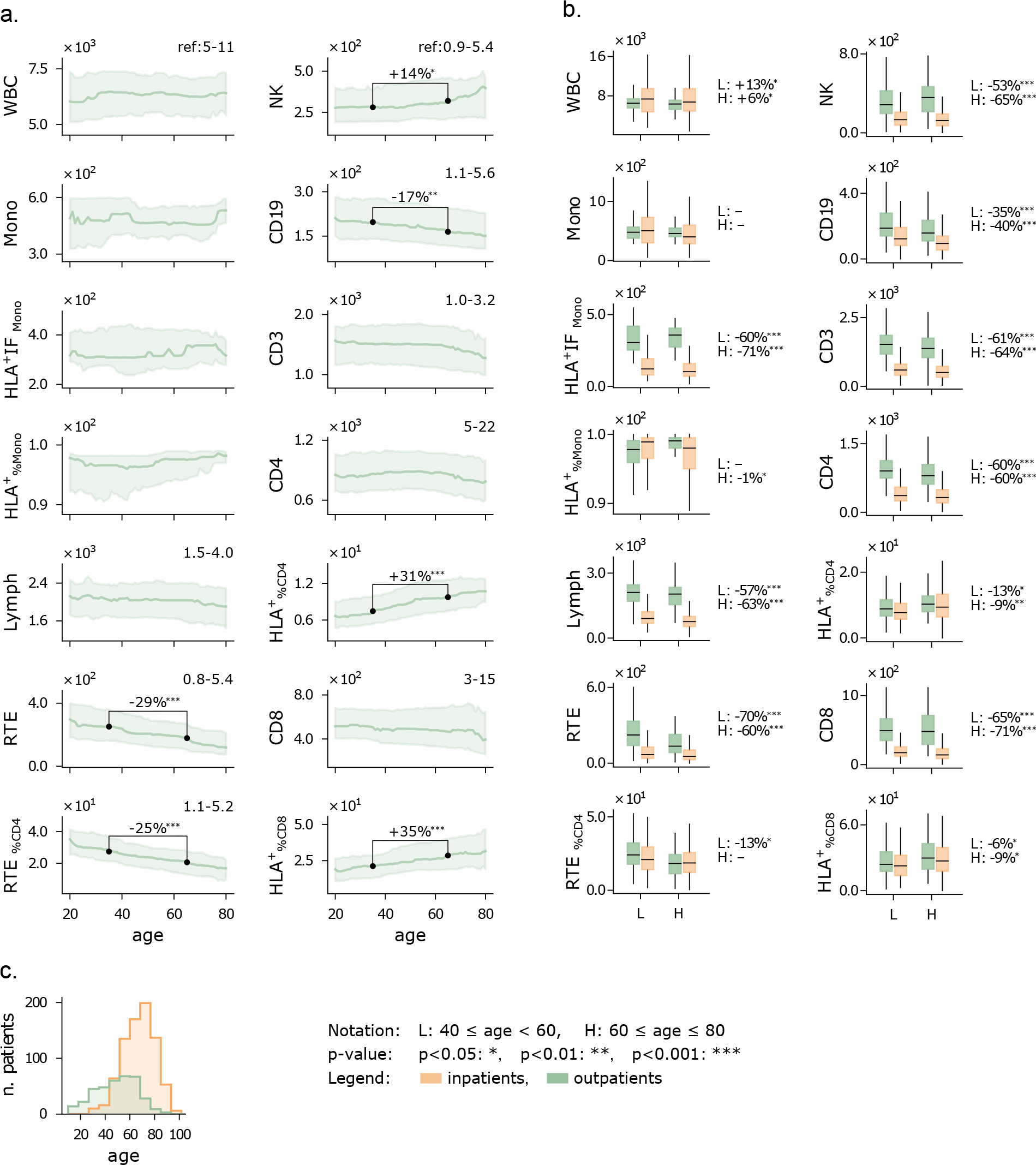
Flow cytometry features of outpatients vs age. **a**) Running median (15-year half-window) of flow cytometry variables. The shaded area is comprised between the first and third running quartiles. The indicated percentage shows a significant increase/decrease of the median at age 65 compared to age 35. **b**) Distribution of flow cytometry variables stratified by age and cohort (inpatients and outpatients). Boxes include first to third quartiles, with the horizontal bar showing the median. The median comparisons shown on the right of each plot are quantified as %-change between the right and left cohorts. **d**) Age histograms of inpatients and outpatients. WBC and related subpopulations are expressed in U*/µ*L.

**Figure 8:**
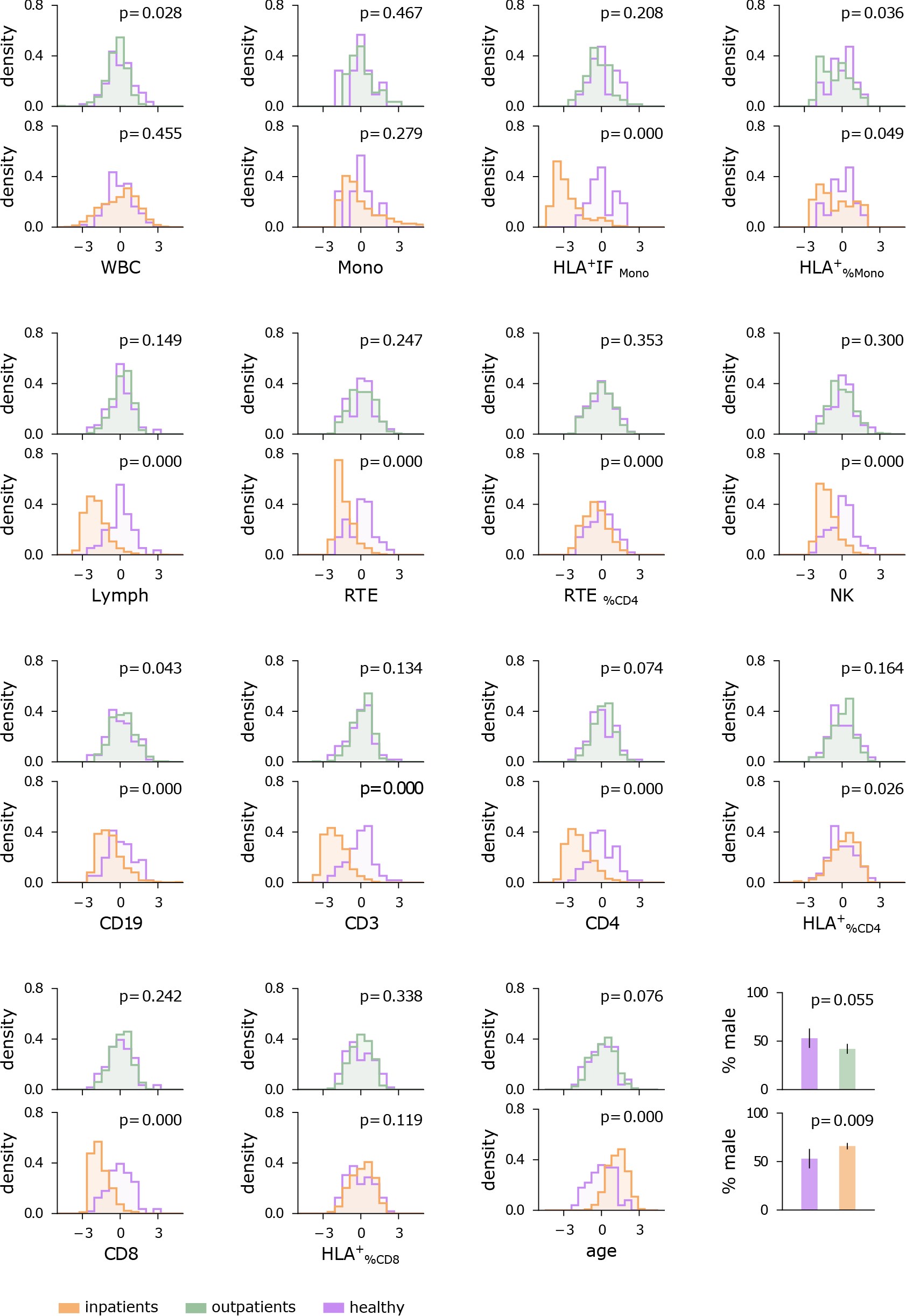
Feature distributions of inpatients, outpatients, and healthy cohort. To ensure comparability, data for all variables were scaled and shifted to standardized statistics within the healthy population. Statistical comparisons between the healthy cohort and outpatients/inpatients were conducted using the Mann–Whitney U test. Proportions of male patients were compared using the z-test. The statistical significance of each comparison is provided by the p-value.

**Table 4:** Extended demographics and clinical characteristics, of the hospitalized patients grouped by age, number of days elapsed between symptoms onset and hospitalization (Δ*t*_ons_), and WHO level, and of the outpatients. Numerical variables (age, CCI, immune cells, cytokines, biomarkers) are expressed in terms of median (Q2), first (Q1) and third (Q3) quartiles, with the format Q2(Q1-Q3). **Demographics:** number of records (Number), age, and proportion of male patients. **Co-morbidities:** proportion of patients with obesity, hypertension, cardiovascular disorders (CVDs), dyslipidemia, diabetes, chronic kidney injuries (CKI), solid and hematologic neoplasms (Tumor, Oncohematology), chronic obstructive pulmonary disease (COPD), autoimmunity, hepatopathy, immunosuppression (primary or secondary). Co-morbidities are also summarized in the Charlson Comorbidity Index (CCI). Immune cells measured via flow cytometry: white blood cells (WBC), monocytes (Mono), mean intensity fluorescence of HLA-DR-positive monocytes (HLA^+^IF Mono), percentage of HLA-DR-positive monocytes (HLA^+^ % Mono), lymphocytes (Lymph), T CD3^+^ cells (CD3), percentage of CD3^+^ lymphocytes (CD3 % Lymph), HLA-DR-positive T CD3^+^ cells (CD3 HLA^+^), Th CD3^+^CD4^+^ cells (CD4), percentage of CD3^+^CD4^+^ lymphocytes (CD4 % Lymph), percentage of HLA-DR-positive CD4 cells (HLA^+^ % CD4), recent thymic emigrants (RTE) and associated percentage of CD4 cells (RTE % CD4), Tc CD3^+^CD8^+^ cells (CD8), percentage of CD3^+^CD8^+^ lymphocytes (CD8 % Lymph), percentage of HLA-DR-positive CD8 cells (HLA^+^ % CD8), B CD3–CD19^+^ cells (CD19), percentage of CD3–CD19^+^ lymphocytes (CD8 % Lymph), T NK CD3–CD56/CD16^+^ cells (NK), and percentage of CD3–CD56/CD16^+^ lymphocytes (NK % Lymph). Cytokines: interferon-γ (INF-γ), interleukin 10 (IL10), IL1-Beta (IL1B), sIL2R-α/sCD25 (IL2R), interleukin 6 (IL6), interleukin 8 (IL8), and interleukin IP10/CXCL10 (IP10). **Biomarkers:** pro-adrenomedullin (proADM), lactate dehydrogenase (LDH), and C-reactive protein (CRP). **Outcomes:** proportion of patients who developed infectious complications, underwent orotracheal intubation (OTI), died (death), or experienced either OTI or death (death^+^OTI). A dash symbol indicates missing data.

| Demographics | age < 70 | age ≥ 70 | Δt<sub>ons</sub> ≤ 10 | Δt<sub>ons</sub> > 10 | WHO ≤ 2 | WHO ≥ 3 | All | Outpatients |
| --- | --- | --- | --- | --- | --- | --- | --- | --- |
| Number | 433 | 357 | 441 | 263 | 263 | 452 | 790 | 367 |
| Age | 60 (54-65) | 78 (74-82) | 67 (58-76) | 69 (59-75) | 66 (56-76) | 69 (60-77) | 68 (59-77) | 51 (36-61) |
| Male | 311/433 (72%) | 212/357 (59%) | 295/441 (67%) | 179/263 (68%) | 148/263 (56%) | 318/452 (70%) | 523/790 (66%) | 153/367 (42%) |

| Co-morbidities | age < 70 | age ≥ 70 | Δt<sub>ons</sub> ≤ 10 | Δt<sub>ons</sub> > 10 | WHO ≤ 2 | WHO ≥ 3 | All | Outpatients |
| --- | --- | --- | --- | --- | --- | --- | --- | --- |
| Obesity | 230/364 (63%) | 143/269 (53%) | 196/343 (57%) | 144/228 (63%) | 127/217 (59%) | 226/356 (63%) | 373/633 (59%) | - |
| Hypertension | 169/428 (39%) | 234/351 (67%) | 227/433 (52%) | 134/261 (51%) | 119/257 (46%) | 241/448 (54%) | 403/779 (52%) | - |
| CVDs | 90/427 (21%) | 156/352 (44%) | 145/434 (33%) | 65/261 (25%) | 84/261 (32%) | 139/444 (31%) | 246/779 (32%) | - |
| Dyslipidemia | 76/400 (19%) | 95/326 (29%) | 92/403 (23%) | 60/245 (24%) | 54/244 (22%) | 102/408 (25%) | 171/726 (24%) | - |
| Diabetes | 62/427 (15%) | 71/353 (20%) | 72/437 (16%) | 45/260 (17%) | 40/260 (15%) | 77/446 (17%) | 133/780 (17%) | - |
| CKI | 25/430 (6%) | 28/354 (8%) | 31/437 (7%) | 15/262 (6%) | 20/261 (8%) | 31/449 (7%) | 53/784 (7%) | - |
| Tumor | 14/427 (3%) | 18/354 (5%) | 19/434 (4%) | 8/261 (3%) | 13/260 (5%) | 17/447 (4%) | 32/781 (4%) | - |
| Oncohematology | 16/429 (4%) | 14/353 (4%) | 16/434 (4%) | 9/262 (3%) | 11/261 (4%) | 18/448 (4%) | 30/782 (4%) | - |
| COPD | 13/425 (3%) | 28/350 (8%) | 28/433 (6%) | 10/257 (4%) | 9/259 (3%) | 28/444 (6%) | 41/775 (5%) | - |
| Autoimmunity | 18/427 (4%) | 23/353 (7%) | 25/434 (6%) | 12/260 (5%) | 16/259 (6%) | 21/447 (5%) | 41/780 (5%) | - |
| Hepatopathy | 24/430 (6%) | 9/355 (3%) | 21/437 (5%) | 9/262 (3%) | 12/262 (5%) | 17/449 (4%) | 33/785 (4%) | - |
| Immunosuppression | 18/428 (4%) | 12/353 (3%) | 19/435 (4%) | 8/261 (3%) | 10/258 (4%) | 16/449 (4%) | 30/781 (4%) | - |
| CCI | 2 (1-3) | 4 (3-5) | 3 (2-4) | 3 (2-4) | 3 (1-4) | 3 (2-4) | 3 (2-4) | - |

| Immune cells | age < 70 | age ≥ 70 | Δt<sub>ons</sub> ≤ 10 | Δt<sub>ons</sub> > 10 | WHO ≤ 2 | WHO ≥ 3 | All | Outpatients |
| --- | --- | --- | --- | --- | --- | --- | --- | --- |
| WBC (10<sup>3</sup> U/μL) | 7.0 (4.8-9.6) | 6.4 (4.8-9.5) | 6.2 (4.2-8.6) | 8.0 (5.9-10.4) | 6.1 (4.1-8.5) | 7.7 (5.4-10.4) | 6.8 (4.8-9.5) | 6.4 (5.3-7.3) |
| Mono (10<sup>2</sup> U/μL) | 4.6 (2.9-7.1) | 3.9 (2.8-5.8) | 3.8 (2.5-5.7) | 5.1 (3.3-7.0) | 4.3 (2.9-6.7) | 4.3 (2.9-6.4) | 4.3 (2.9-6.3) | 4.7 (3.9-5.9) |
| HLA<sup>+</sup> IF Mono (10<sup>2</sup> U) | 1.1 (0.8-1.7) | 1.0 (0.7-1.6) | 1.1 (0.8-1.8) | 0.9 (0.7-1.4) | 1.4 (0.8-2.2) | 0.9 (0.6-1.3) | 1.1 (0.7-1.7) | 3.2 (2.7-4.2) |
| HLA<sup>+</sup> % Mono | 98.7 (96.0-99.5) | 98.0 (94.0-99.4) | 98.7 (96.0-99.6) | 98.0 (93.7-99.1) | 99.0 (97.0-99.7) | 98.0 (94.0-99.3) | 98.4 (95.0-99.5) | 98.7 (96.5-99.2) |
| Lymph (10<sup>2</sup> U/μL) | 8.5 (6.1-11.5) | 7.0 (4.8-9.4) | 7.9 (5.4-10.1) | 7.2 (5.0-10.3) | 9.0 (6.5-11.9) | 7.0 (5.0-9.8) | 7.9 (5.5-10.5) | 20.4 (16.5-24.0) |
| CD3 (10<sup>2</sup> U/μL) | 5.6 (3.8-8.1) | 4.6 (2.9-6.8) | 5.1 (3.3-7.1) | 4.9 (3.1-7.3) | 6.0 (4.3-8.6) | 4.6 (2.9-6.7) | 5.1 (3.3-7.4) | 15.0 (11.4-18.2) |
| CD3 % Lymph | 68.0 (59.0-76.0) | 67.0 (57.0-75.0) | 67.0 (57.0-75.0) | 66.0 (58.0-76.0) | 70.0 (62.0-77.0) | 65.0 (56.0-74.0) | 68.0 (58.0-76.0) | 74.0 (67.0-79.8) |
| CD3 HLA<sup>+</sup> (10<sup>1</sup> U/μL) | 7.2 (4.2-11.8) | 6.8 (3.8-11.7) | 6.7 (3.7-10.4) | 6.4 (3.9-11.6) | 8.6 (4.9-14.4) | 6.3 (3.6-9.8) | 7.1 (4.0-11.8) | 20.1 (13.4-30.4) |
| CD4 (10<sup>2</sup> U/μL) | 3.6 (2.4-5.4) | 2.9 (1.8-4.4) | 3.2 (2.1-4.7) | 3.1 (2.0-5.1) | 3.9 (2.5-5.9) | 2.9 (1.9-4.5) | 3.3 (2.1-5.1) | 8.6 (6.8-10.7) |
| CD4 % Lymph | 43.0 (35.0-52.0) | 43.0 (33.0-51.0) | 42.0 (33.0-51.0) | 44.0 (35.0-52.0) | 45.0 (36.0-54.0) | 42.0 (33.0-50.2) | 43.0 (34.0-51.8) | 44.0 (39.0-49.0) |
| HLA<sup>+</sup> % CD4 | 8.3 (5.9-12.2) | 10.3 (6.8-14.6) | 8.6 (5.9-12.9) | 9.8 (6.7-13.6) | 9.4 (6.0-13.0) | 9.4 (6.5-13.6) | 9.3 (6.4-13.3) | 8.7 (6.3-11.5) |
| RTE (10<sup>1</sup> U/μL) | 6.7 (3.6-12.0) | 4.5 (2.1-8.3) | 5.3 (2.5-10.1) | 5.5 (3.2-10.6) | 6.9 (3.4-12.4) | 5.1 (2.7-9.7) | 5.5 (2.9-10.5) | 20.7 (12.7-32.9) |
| RTE % CD4 | 20.9 (14.2-28.3) | 17.4 (9.9-23.8) | 18.4 (12.2-26.1) | 20.2 (12.1-28.9) | 19.0 (13.0-27.4) | 19.1 (12.9-26.5) | 19.0 (12.3-26.7) | 24.4 (17.1-33.2) |
| CD8 (10<sup>2</sup> U/μL) | 1.6 (1.1-2.5) | 1.3 (0.7-2.2) | 1.4 (0.9-2.4) | 1.4 (0.9-2.1) | 1.8 (1.1-2.8) | 1.3 (0.8-2.1) | 1.5 (0.9-2.4) | 4.9 (3.5-6.7) |
| CD8 % Lymph | 21.0 (14.0-26.0) | 18.0 (13.0-27.0) | 20.0 (14.0-26.0) | 19.0 (13.0-25.0) | 21.0 (14.5-27.5) | 19.0 (13.0-25.0) | 20.0 (14.0-26.0) | 25.0 (20.0-31.0) |
| HLA<sup>+</sup> % CD8 | 23.1 (14.3-33.3) | 31.0 (19.2-44.4) | 25.0 (15.4-36.3) | 27.3 (16.1-40.0) | 25.0 (17.1-36.8) | 26.3 (16.6-38.1) | 26.3 (16.7-38.5) | 23.8 (16.1-35.6) |
| CD19 (10<sup>1</sup> U/μL) | 11.5 (7.5-18.4) | 7.2 (4.3-11.6) | 9.2 (5.4-14.4) | 10.7 (6.3-15.7) | 9.9 (5.5-15.3) | 9.9 (5.9-15.2) | 9.8 (5.5-14.7) | 18.8 (13.0-26.3) |
| CD19 % Lymph | 15.0 (10.0-20.0) | 11.0 (7.0-17.0) | 12.0 (8.0-18.0) | 15.0 (11.0-21.0) | 11.0 (7.0-17.0) | 15.0 (10.0-20.5) | 13.0 (9.0-19.0) | 9.4 (7.0-12.0) |
| NK (10<sup>2</sup> U/μL) | 1.3 (0.8-2.0) | 1.2 (0.7-2.0) | 1.3 (0.8-2.1) | 1.1 (0.6-1.8) | 1.5 (0.8-2.1) | 1.2 (0.7-1.8) | 1.3 (0.7-2.0) | 3.0 (2.0-4.4) |
| NK % Lymph | 15.0 (10.0-23.0) | 18.4 (11.0-28.0) | 18.0 (12.0-27.0) | 15.0 (9.0-23.0) | 16.0 (10.0-24.0) | 16.0 (10.0-26.0) | 17.0 (10.0-25.0) | 15.0 (11.0-22.0) |

| Cytokines | age < 70 | age ≥ 70 | Δt<sub>ons</sub> ≤ 10 | Δt<sub>ons</sub> > 10 | WHO ≤ 2 | WHO ≥ 3 | All | Outpatients |
| --- | --- | --- | --- | --- | --- | --- | --- | --- |
| IFN-γ (pg/mL) | 1.7 (0.4-5.4) | 1.6 (0.3-4.5) | 2.1 (0.5-5.7) | 1.4 (0.2-4.3) | 1.1 (0.2-4.2) | 1.9 (0.5-5.1) | 1.7 (0.3-5.0) | - |
| IL10 (10<sup>1</sup> pg/mL) | 1.2 (0.7-2.2) | 1.5 (0.8-2.2) | 1.6 (0.9-2.3) | 1.1 (0.7-2.0) | 0.9 (0.6-1.6) | 1.7 (0.9-2.5) | 1.4 (0.7-2.2) | - |
| IL1B (pg/mL) | 7.7 (0.9-16.6) | 10.7 (1.6-18.8) | 10.9 (3.9-21.5) | 7.5 (0.3-15.4) | 6.3 (0.3-10.6) | 9.8 (0.3-20.2) | 8.5 (0.9-18.5) | - |
| IL2R (10<sup>3</sup> pg/mL) | 2.9 (2.2-3.9) | 3.5 (2.6-4.5) | 3.0 (2.3-4.0) | 3.4 (2.6-4.5) | 2.9 (2.1-4.1) | 3.4 (2.6-4.5) | 3.2 (2.3-4.2) | - |
| IL6 (10<sup>1</sup> pg/mL) | 2.9 (1.2-5.8) | 3.3 (1.9-7.2) | 3.4 (1.7-6.7) | 2.7 (1.2-5.4) | 2.1 (0.9-4.3) | 3.6 (1.9-8.2) | 3.0 (1.4-6.3) | - |
| IL8 (10<sup>1</sup> pg/mL) | 3.3 (2.2-5.2) | 4.1 (3.0-6.2) | 3.8 (2.6-5.9) | 3.4 (2.3-5.1) | 3.1 (2.0-4.4) | 4.2 (2.7-6.3) | 3.6 (2.6-5.7) | - |
| IP10 (10<sup>3</sup> pg/mL) | 1.1 (0.6-1.7) | 1.4 (1.0-1.8) | 1.4 (0.9-2.0) | 1.1 (0.6-1.6) | 0.9 (0.4-1.4) | 1.5 (1.0-2.0) | 1.2 (0.7-1.8) | - |

| Biomarkers | age < 70 | age ≥ 70 | Δt<sub>ons</sub> ≤ 10 | Δt<sub>ons</sub> > 10 | WHO ≤ 2 | WHO ≥ 3 | All | Outpatients |
| --- | --- | --- | --- | --- | --- | --- | --- | --- |
| proADM (nmol/L) | 0.8 (0.7-1.1) | 1.2 (1.0-1.6) | 1.0 (0.8-1.4) | 0.9 (0.7-1.2) | 0.9 (0.7-1.3) | 1.1 (0.8-1.4) | 1.0 (0.8-1.3) | - |
| LDH (10<sup>2</sup> U/L) | 5.8 (4.4-7.5) | 6.2 (4.5-7.6) | 5.8 (4.4-7.3) | 6.6 (5.1-7.9) | 4.8 (3.8-6.4) | 6.8 (5.4-8.4) | 6.0 (4.5-7.6) | - |
| CRP (10<sup>1</sup> mg/L) | 6.9 (3.1-10.5) | 7.0 (3.4-11.3) | 7.1 (3.4-11.0) | 7.0 (3.7-11.3) | 4.7 (1.9-9.9) | 8.1 (4.5-12.2) | 7.0 (3.3-11.0) | - |

Table 4: *Extended demographics and clinical characteristics*, of the hospitalized patients grouped by age, number of days elapsed between symptoms onset and hospitalization ( $\Delta t_{\text{ons}}$ ), and WHO level, and of the outpatients.
| Outcomes | age < 70 | age ≥ 70 | Δt<sub>ons</sub> ≤ 10 | Δt<sub>ons</sub> > 10 | WHO ≤ 2 | WHO ≥ 3 | All | Outpatients |
| --- | --- | --- | --- | --- | --- | --- | --- | --- |
| Infectious complications | 42/433 (10%) | 43/356 (12%) | 58/441 (13%) | 17/262 (6%) | 20/263 (8%) | 57/451 (13%) | 85/789 (11%) | - |
| OTI | 60/430 (14%) | 59/355 (17%) | 70/438 (16%) | 42/262 (16%) | 1/262 (0%) | 105/448 (23%) | 119/785 (15%) | - |
| Death | 39/433 (9%) | 99/357 (28%) | 83/441 (19%) | 35/263 (13%) | 13/263 (5%) | 114/452 (25%) | 138/790 (17%) | - |
| death+OTI | 72/433 (17%) | 118/357 (33%) | 113/441 (26%) | 52/263 (20%) | 13/263 (5%) | 157/452 (35%) | 190/790 (24%) | - |

